# Quantifying the impact of bacterial vaccines against antibiotic resistance: accounting for transmission and selection dynamics

**DOI:** 10.64898/2026.08.25.26361172

**Authors:** Chloé Aupépin, Lulla Opatowski, Iris van Bommel, Elske Sieswerda, Valentijn Schweitzer, Stéphane Loisel, Laura Temime, Quentin J Leclerc

## Abstract

Vaccines, by reducing bacterial infection, transmission and/or colonisation, are promising investments against the global rise of antibiotic resistance (ABR). From a public health perspective, while efforts are put in developing bacterial vaccines, anticipating their potential impact on ABR is essential.

We developed a compartmental model formalising inter-individual transmis-sion and selection pressure through both bystander and targeted antibiotic exposure. Following a mathematical analysis of the model’s equilibrium points, we explored the impact of different vaccines through simulations for two bacterial types.

In simulations, vaccines consistently reduced infection incidence, although to varying extents. For *S. aureus*, a vaccine reducing acquisition rate, infection rate and colonisation duration by 60% at 70% coverage reduced total infections by 80%, while this reduction was only of 48% for *E. coli*. The impact on the resistance proportion among colonised differed markedly: this same vaccine increased it by 11% for *S. aureus*, while decreasing it by 8% for *E. coli*.

Overall, our results highlight that population level impact on ABR strongly depends on the vaccine mechanism of action. The proposed model, which gathers the main drivers involved, provides a general framework that can be adapted to a wide range of bacterial pathogens and vaccines.

## 1 Introduction

Antibiotic resistance (ABR) is increasingly causing standard antibiotics to be ineffective, leading to longer infections and worse health outcomes. ABR represents a major rising public health burden: in 2019, an estimated 5 million deaths worldwide were associated with bacterial antibiotic resistance [1].

ABR operates across multiple scales. When someone is colonised or infected by both resistant and sensitive bacteria, antibiotic exposure can exert a selective pressure, leading to sensitive bacteria being eliminated, while resistant strains survive and become dominant. For a given bacterial species, antibiotic exposure can occur in two ways: directly or indirectly. A direct antibiotic exposure corresponds to treatment administered for an infection caused by that bacterial species. An indirect exposure, which we refer to as bystander exposure, corresponds to a situation where the species of interest is only colonising the host, while antibiotics are administered to treat an infection caused by another pathogen. Both direct and bystander exposures impose selective pressures that can promote the survival and expansion of resistant strains within hosts, thereby contributing to increased resistance prevalence [2]. As individuals carrying bacteria transmit them to others, resistance can spread at the population level, contributing to the broader public health challenge [3].

Given the alarming increase in ABR, preventive strategies - such as vaccination - may have an essential role to play. Vaccines may limit the burden associated with ABR through multiple complementary mechanisms [4, 5]. First, they can directly prevent certain bacterial infections. For example, vaccines against *Streptococcus pneumoniae* and *Haemophilus influenzae* type b have significantly reduced the incidence of invasive infections, such as pneumonia and meningitis, which account for a large proportion of antibiotic use in the population due to high frequency of occurence [6, 7]. By lowering the incidence of all infections, vaccines directly reduce the incidence of antibiotic-resistant infections, but also, reduce the need for antibiotic use and, consequently, the selective pressure which benefits resistant bacteria. Second, vaccines may also reduce colonisation prevalence. For instance, pneumococcal conjugate vaccines induce a substantial decline in the colonisation by the *S. pneumoniae* serotypes they target. This occurs when vaccine-induced antibodies either decrease the risk of acquisition or accelerate decolonisation. Since individuals are more at risk of developing infections from bacteria they already carry, reducing the number of colonised individuals leads to a decrease in the incidence of infections. Additionally, lowering the colonisation prevalence of all strains reduces the circulation of pathogens within the population and slows down the spread of all bacterial strains including the resistant ones.

However, while some vaccines targeting bacterial pathogens are available and several are under development, the population-level impact of those on ABR remains insufficiently understood [8]. Moreover, regular clinical studies like trials or observational cohorts take time, may include biases impeding generalisation to the entire population and would not be able to quantify indirect impacts [9]. In this context, mathematical modelling is a valuable tool to understand this impact and guide the development of new vaccines while accounting for the complex resistance transmission and selection dynamics described above [10]. For instance, over the last 20 years, several studies have examined the introduction of pneumococcal conjugate vaccines to assess their population-level effects and the emergence of serotype replacement. Some of this work suggested that vaccination alone may not be sufficient to control the spread of resistant strains [11], while other studies found that reduced antibiotic use, combined with serotype replacement, may lead to increased infections by antibiotic-sensitive strains after vaccine implementation [12]. Additional research explored the combined impact of vaccines and antibiotics, including the hypothetical introduction of novel antibiotics in vaccinated populations [13]. There is also growing interest in modelling the impact of hypothetical vaccines, such as those targeting *Staphylococcus aureus* [14, 15] *or Mycobacterium tuberculosis* [16, 17], to assess their potential benefits, guide their development and compare them to other interventions.

Nevertheless, these previous studies have, for the most part, provided specific models to focus on single, specific pathogens. This focus cannot provide global view of the potential impact of vaccines to support guidance for policy decisions aiming to tackle the global ABR crisis, as resistance is a multifactorial issue involving numerous bacteria and antibiotics. Pathogen-specific studies are often difficult to compare. To address this gap, the World Health Organization (WHO) has developed a unified modelling framework to estimate the impact of vaccines on ABR, enabling comparison between vaccines and identifying those with the greatest potential impact. However, the WHO framework is static: it does not account for transmission and selection dynamics of bacteria or indirect vaccine impact on non-vaccinated individuals through herd immunity [18], which therefore limits the evaluation of real vaccine impacts in the population.

In this study, we propose a generic modelling framework of bacterial transmission dynamics modulated by the use of vaccines, that enables the exploration of a wide range of epidemiological contexts and potential outcomes like incidence of resistant infections or proportion of resistance among colonised individuals, offering a more comprehensive understanding of how vaccination may influence ABR.

## 2 Methods

### 2.1 Model structure

We developed a generic compartmental deterministic model to represent the dynamics of colonisation and infection by bacteria in the community. The model structure includes differences between antibiotic-sensitive and antibiotic-resistant strain and incorporates both direct and bystander antibiotic exposure and vaccination status (Figure 1).

**Figure 1:**
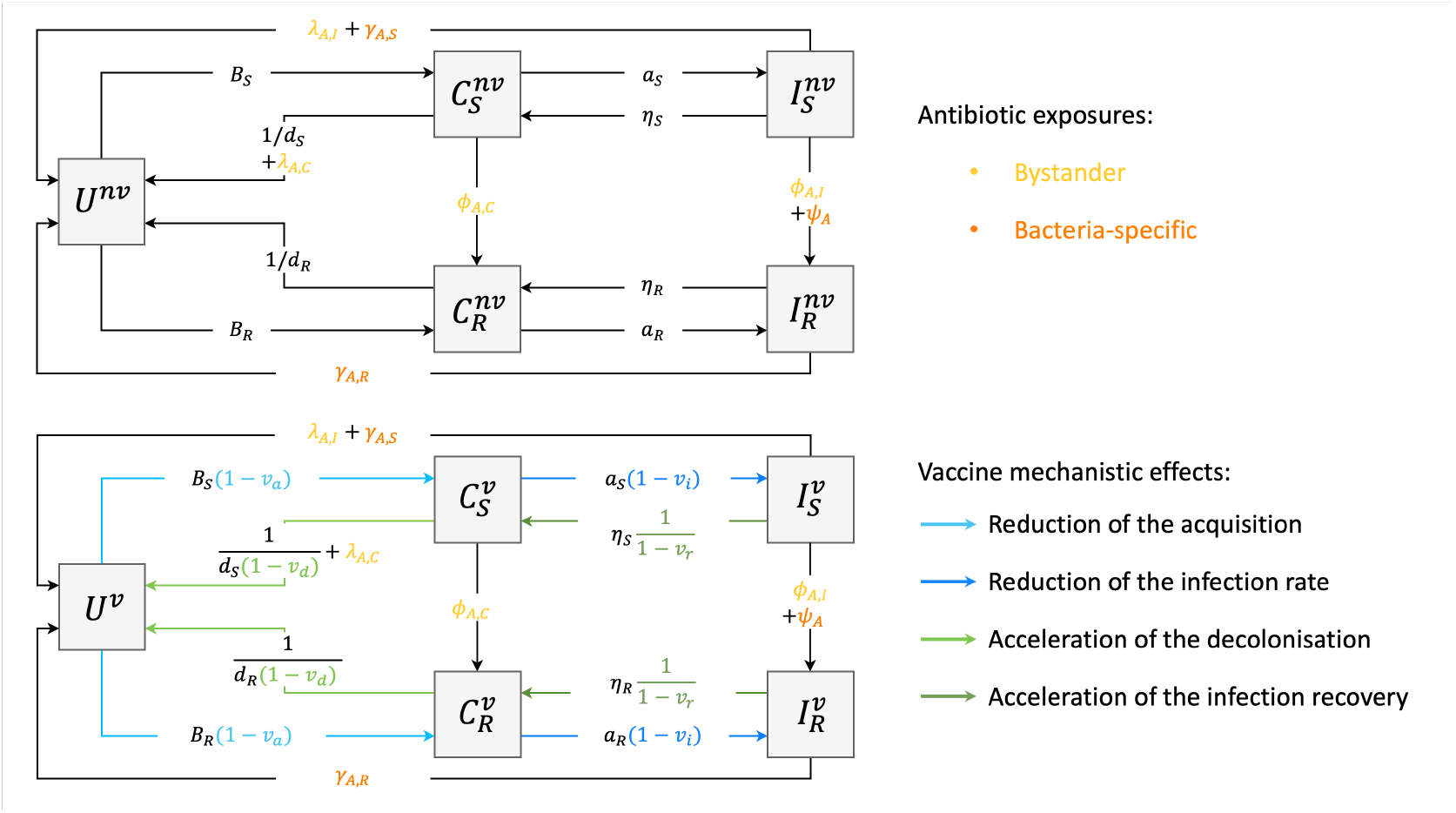
Model structure. Individuals can be uncolonised (U), colonised (C) or infected (I). Colonisation or infection can be by either an antibiotic-sensitive or resistant strain (subscript S or R). Individuals are vaccinated or not (superscripts *v* or *nv*). Both colonised and infected individuals can transmit bacterial strains, with a reduced transmission strength for resistant bacteria modelled considering a relative fitness cost *f*. There are 4 possible effects of a vaccine: reduction of acquisition (*v*_*a*_), reduction of infection (*v*_*i*_), acceleration of decolonisation (*v*_*d*_), and acceleration of infection recovery (*v*_*r*_). Yellow parameters refer to bystander antibiotic exposure. Orange parameters refer to antibiotic exposure specific to an infection caused by the modelled bacteria.

#### 2.1.1 Bacterial colonisation

The population is divided into three main categories: uncolonised individuals, colonised individuals, and infected individuals.

##### Transmission

Uncolonised individuals can become colonised by either antibiotic-sensitive or antibiotic-resistant bacteria upon contact with colonised or infected individuals. Let *U* denote uncolonised individuals, *C*_*S*_ and *C*_*R*_ the colonised individuals with sensitive and resistant strains, respectively, and *I*_*S*_ and *I*_*R*_ the corresponding infected individuals. Transmission occurs via contact with colonised or infected hosts, with rates *B*_*S*_ and *B*_*R*_ defined as

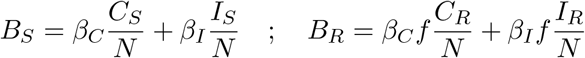

where *β*_*C*_ and *β*_*I*_ are the transmission rates from colonised and infected individuals, and *N* is the total population size. We can assume *β*_*I*_ ≤ *β*_*C*_ if infected individuals are hospitalised or self-isolating to some extent, thereby reducing their contacts. Conversely, *β*_*I*_ ≥ *β*_*C*_ may hold if infected individuals maintain their contacts, but with a higher bacterial load, leading to greater transmissibility. To differentiate transmission of sensitive and resistant bacteria, we used a relative fitness parameter, ranging from 0 to 1, acting as a multiplicative factor. This parameter is commonly used in mathematical models to represent the cost of antibiotic resistance mechanisms development from the bacterial perspective [10].

##### Natural clearance

Most bacteria colonise their human hosts for a given colonisation duration, denoted *d*_*S*_ for the sensitive strain and *d*_*R*_ for the resistant strain. After this, colonised individuals naturally return to an uncolonised state through spontaneous bacterial clearance.

##### Bystander antibiotic exposure

In addition to the specifically modelled bacteria, individuals may be infected by another pathogen for which they are prescribed antibiotics. This situation is referred to as bystander exposure. We suppose that a fixed proportion (denoted *p*_*by*_) of individuals are subject to bystander exposure, and that antibiotics clear the sensitive bacteria within a time *τ*_*dec,by*_. Furthermore, in individuals colonised by a sensitive strain, by-stander exposure can select for a pre-existing minority resistant strain with probability *p*_*min,C*_. As a result, bystander exposure has two possible effects:

i. decolonisation of sensitive bacteria, occurring at rate 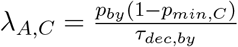 and
ii. selection of a resistant strain, occurring at rate 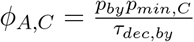.

This bystander exposure also occurs in infected individuals in the model, as described below.

#### 2.1.2 Bacterial infection

A colonised individual may develop an infection at a rate *a*_*S*_ for sensitive strains and *a*_*R*_ for resistant strains. We assume that a fixed proportion of individuals infected by a sensitive strain are prescribed antibiotics because of the infection (*p*_*sp,S*_) and that these antibiotics clear the bacteria in a certain amount of time (*τ*_*dec,sp,S*_), reverting individuals to the uncolonised state. Alternatively, direct antibiotic exposure can select for a minority resistant strain present in a fraction of individuals infected by sensitive bacteria (*p*_*min,I*_). This second action implies that the antibiotic taken to treat the infection can select the studied resistance. Thus, the decolonisation rate is

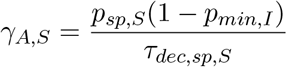

and the selection rate is

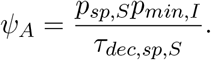

A fraction 1 − *p*_*sp,S*_ of infections do not require antibiotics. In this case, infected individuals may still be exposed to antibiotics through bystander exposure which can decolonise sensitive strains at a rate

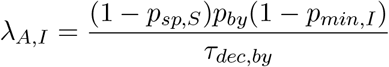

or select resistant strains at a rate

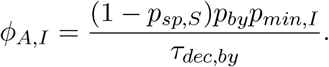

Finally, when no antibiotics are taken, infected individuals may naturally recover and come back to a colonised state at a rate

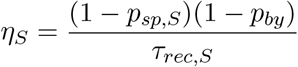

where *τ*_*rec,S*_ represents the time until natural recovery.

Furthermore, we assume that a fraction *p*_*sp,R*_ of individuals infected by a resistant strain will take second-line antibiotics, and that the ensuing decolonisation rate will be

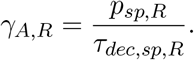

The fraction 1 − *p*_*sp,R*_ that does not take antibiotics will naturally recover at a rate

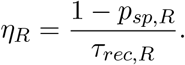

#### 2.1.3 Vaccination

Individuals can either be vaccinated (*v*) or non-vaccinated (*nv*). Vaccination coverage is defined in the initial conditions, and individuals do not transition between non-vaccinated and vaccinated states over time. Vaccinated and non-vaccinated individuals interact through transmission: a vaccinated individual can transmit bacteria to a non-vaccinated individual and vice-versa.

The vaccine has 4 possible effects, each with an efficacy ranging between 0 and 100%. It can reduce acquisition with an efficacy *v*_*a*_, whereby the transmission rate is multiplied by 1 − *v*_*a*_. It can accelerate decolonisation with an efficacy *v*_*d*_, whereby the decolonisation rate is multiplied by 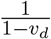. It can reduce the risk of infection with an efficacy *v*_*i*_, whereby the infection rate is multiplied by 1 − *v*_*i*_. Finally, it can accelerate recovery with an efficacy *v*_*r*_, whereby the recovery rate is multiplied by 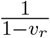. We built types of vaccines by combining these four effects; a vaccine could act on one, two, three, or all four of these effects, resulting in 15 possible vaccine types presented in Table 1.

**Table 1:** List of vaccine types. Fifteen different vaccine types were considered, combining four types of effects. A cross in a cell indicates that the type of vaccine includes the effect presented in the column. Vaccines reducing acquisition are denoted with subscript a, vaccines accelerating acquisition with subscript d, vaccines reducing the infection rate with subscript i, and vaccines accelerating infection recovery with subscript r.

| Symbol | Reduce acquisition | Accelerate decolonisation | Reduce infection rate | Accelerate infection recovery |
| --- | --- | --- | --- | --- |
| $V_a$ | x | | | |
| $V_d$ | | x | | |
| $V_i$ | | | x | |
| $V_r$ | | | | x |
| $V_{a,d}$ | x | x | | |
| $V_{a,i}$ | x | | x | |
| $V_{a,r}$ | x | | | x |
| $V_{d,i}$ | | x | x | |
| $V_{i,r}$ | | | x | x |
| $V_{d,r}$ | | x | | x |
| $V_{a,d,i}$ | x | x | x | |
| $V_{a,d,r}$ | x | x | | x |
| $V_{a,i,r}$ | x | | x | x |
| $V_{d,i,r}$ | | x | x | x |
| $V_{a,d,i,r}$ | x | x | x | x |

Different levels of vaccine coverage (10%, 30%, 50%, 70% and 90%) and vaccine efficacy parameter values (0.3, 0.6 and 0.9) were explored to assess their impact. When the vaccine combined several effects, all efficacy values were assumed equal.

#### 2.1.4 Model equations

Changes over time in the number of individuals in each model compartment are driven by the following set of ordinary differential equations:

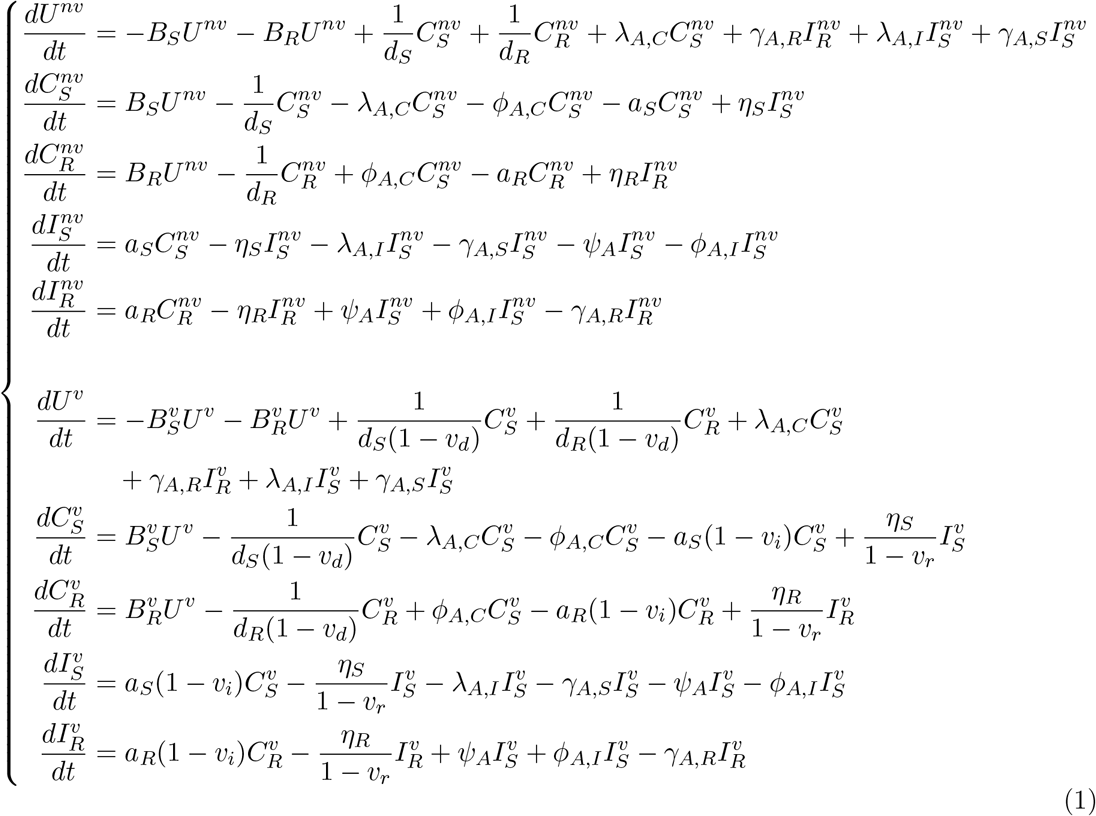

where 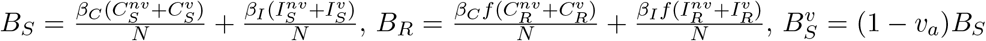 and 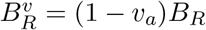.

Throughout this work, we often assume identical bacterial characteristics for both sensitive and resistant strains, in which case we remove the subscripts S or R from parameter notation.

### 2.2 Equilibria analyses

For many bacterial species, antibiotic-sensitive and antibiotic-resistant strains coexist at the population level [19], as observed for instance in *Streptococcus pneumoniae* [20] and *Escherichia coli* [21]. Thus, knowing under which conditions a steady-state with coexistence is allowed by the model structure is important. To address this question, we analytically derived equilibria under a range of simplifying assumptions summarised in Table 2.

**Table 2:** List of assumptions for equilibria analysis. Different combinations of antibiotic-induced model transitions were considered. When “No” is written, the corresponding flux was set to zero. Columns 3 and 5 correspond to decolonisation by antibiotic exposure. Columns 4, 6 and 7 correspond to selection of resistant strain by antibiotic exposure.

| n° | Vaccine coverage | Antibiotic $C_S \rightarrow U$ | Antibiotic $C_S \rightarrow C_R$ | Antibiotic $I_S \rightarrow U$ | Antibiotic $I_S \rightarrow I_R$ | Antibiotic $I_R \rightarrow U$ |
| --- | --- | --- | --- | --- | --- | --- |
| 1 | 0% | No | No | No | No | No |
| 2 | 0% | Yes | No | Yes | No | Yes |
| 3 | 0% | Yes | Yes | Yes | Yes | Yes |
| 4 | 100% | No | No | No | No | No |
| 5 | 100% | Yes | No | Yes | No | Yes |
| 6 | 100% | Yes | Yes | Yes | Yes | Yes |

We computed basic reproduction numbers, which characterise the intrinsic transmissibility of strains according to their epidemiological and biological features, to summarise the essential mechanisms of bacterial transmission and evaluate the impact of vaccines on these indicators. They were analytically derived for both acquisition of the sensitive and resistant strains in the absence of the other strain (see Appendix A.1).

### 2.3 Numerically identifying key parameters for bacterial dynamics and vaccine impact

To identify key parameters for bacterial dynamics and vaccine impact, we conducted two separate analyses. Within these analyses we explored a range of pathogens.

First, to identify which of the parameters influence bacterial dynamics in the absence of a vaccine, we computed the equilibrium in a fully unvaccinated population. We then simulated one year from this equilibrium point and computed the annual cumulative incidence of infections (total, sensitive, resistant), the proportion of resistant infections, and the prevalence of colonisation (total, sensitive, resistant) with the associated proportion of resistant colonisation.

Then, to identify which of the parameters influence vaccine impact, we again used the equilibrium obtained in a fully unvaccinated population as the initial condition. From this point, we simulated disease dynamics over one year without vaccination and computed the same outputs as before. This scenario was then compared to a one-year simulation with vaccination in place. We assumed that the vaccine was distributed uniformly across all population groups, with immediate efficacy.

In order to represent the diversity in existing bacterial characteristics, parameter ranges were defined based on available literature (Table A1) and are purposefully wide. For both analyses, we explored the parameter space through a Latin Hypercube Sampling design (5 000 samples) combined with Partial Rank Correlation Coefficient (PRCC) computation [22]. Only the parameter sets allowing coexistence between sensitive and resistant strains were kept in the analysis. For the first analysis, we observed the effect of varying parameters on absolute values of the model outputs, for the second one, we observed the effect of varying parameters on the relative vaccination-induced reduction of the model outputs considered.

### 2.4 Case study: application to two specific bacteria

Finally, we explored the potential impacts of vaccination through two specific case studies. We focused on *S. aureus* and *E. coli*, as both are associated with community-acquired infections and are considered high priority pathogens by the WHO [18]. Those bacteria are also particularly relevant due to their differences in incidence of infection in the community, colonisation prevalence and resistance prevalence which prove how the generic model can be applied to various bacteria.

#### 2.4.1 Epidemiology of *Staphylococcus aureus* and case study assumptions

*S. aureus* colonises on average 30% of the population at any given time and the estimated prevalence of methicillin-resistant *S. aureus* (MRSA) is 10% in France [23]. *S. aureus* is responsible for a large variety of infections. For this case study, we focus on bacteraemia. The annual incidence rate of bacteraemia caused by *S. aureus* in Europe was estimated at 37.9 per 100 000 inhabitants in 2024 [23]. The standard treatment for bacteraemia is a 2-week course of intravenous antibiotics such as flucloxacillin for methicillin-sensitive *S. aureus* (MSSA) and vancomycin for MRSA [24]. Given that bacteraemia is a severe infection, we assumed that every infected individual would take antibiotics. The main antibiotics responsible for bystander exposure for *S. aureus* were assumed to be beta-lactamase-resistant penicillins (Oxacillin, Nafcillin, Methicillin, Flucloxacillin) and combinations of penicillins with beta-lactamase inhibitors (Amoxicillin with clavulanic acid) as these are prescribed in the community and effective on MSSA but not on MRSA resulting in resistance selection [25, 26, 27].

#### 2.4.2 Epidemiology of *Escherichia coli* and case study assumptions

*E. coli* colonises more than 90% of the population [28], and in France about 6% of strains are resistant to third-generation cephalosporins [29]. These bacteria are an important cause of all urinary tract infections (UTIs), which we will focus on for this case study. The estimated annual age-standardised incidence rate for UTIs (all pathogens combined) in France was 7 691 per 100 000 inhabitants in 2021, which corresponds to a daily incidence rate of approximately 21 per 100 000 inhabitants [30]. *E. coli* causes around 80% of all UTIs, therefore we chose to fix the daily incidence of *E. coli* UTIs to 16 per 100 000 individuals in our analysis [31]. We consider that 80% of infections are treated by antibiotics, the remaining 20% of the infected individuals do not require antibiotics and will either be indirectly exposed to antibiotics or naturally recover to a colonised state. We assume that 20% of infections are complicated UTIs, using a cross-sectional study in the outpatient setting in France [32]. Thus, 60% of UTI cases take the first-line antibiotic treatment (fosfomycin), which induces bacterial clearance in 2 days irrespective of the cephalosporin resistance status, and does not select for resistant strains. For the 20% of UTIs which are complicated and require alternative treatment: for complicated sensitive UTIs, the standard treatment would be third generation cephalosporins, with a treatment duration of 7 days, and which can select for resistant strains. For complicated resistant UTIs, the standard treatment would be carbapenems, with an average treatment duration of 9 days [33]. This parametrisation leads to the following expressions of antibiotic effect rates:

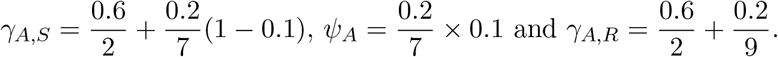

The main antibiotics responsible for bystander exposure selecting for the resistance of *E. coli* against third-generation cephalosporins are third-generation cephalosporins and fluoroquinolones, as the resistance to the latter is present on mobile genetic elements and co-selects for beta-lactam resistance [34].

#### 2.4.3 Model calibration

We calibrated the transmission rate, the relative fitness *f* and the infection rate *a* to reproduce the prevalence of colonisation, the prevalence of resistant strains among carried bacteria and the incidence of infections associated to each bacterium. All three parameters were optimised at the same time by the function nlminb which minimises an objective function using a quasi-Newton optimisation method starting at an initial value and allowing the definition of parameter bounds. A multi-start optimisation approach was used by defining 10 initial values uniformly distributed within bounds for each parameter, leading to 1 000 parameter sets. Among the optimal sets of parameters obtained, the best one was selected as the one giving the lowest value of the objective function.

The objective function was defined as the sum of the squared differences between the three target outputs (prevalence of colonisation, prevalence of resistant strains among carried bacteria and incidence of infections) and the values obtained from the analytical computation of the equilibrium (*C*_*S*_ + *C*_*R*_, *C*_*R*_*/*(*C*_*S*_ + *C*_*R*_) and *a*_*S*_*C*_*S*_ + *a*_*R*_*C*_*R*_).

For the *S. aureus* case study, due to the severity of bacteraemia, infected individuals were assumed to be hospitalised and thus not spreading bacteria in the community, translating to *β*_*I*_ = 0. Conversely, for the *E. coli* case study, due to the high prevalence and low severity of UTIs, we considered that transmission was identical in infected and colonised individuals (*β*_*C*_ = *β*_*I*_ = *β*).

All other parameters were fixed based on literature. Parameter values are reported in Table A2 for the *S. aureus* case study and Table A3 for the *E. coli* case study.

#### 2.4.4 Simulations

For both case studies, 500 sets of parameters were obtained by random sampling from a uniform distribution ranging from 10% below to 10% above the parameter values, in order to account for bacterial variability. All parameters (calibrated and fixed) were varied except the relative fitness, since any small change in this parameter disrupts the coexistence condition and, when coexistence is satisfied, the proportion of resistance is mainly driven by its value (Figure A1, see Results - Conditions for coexistence). We then quantified the impact of the types of vaccines defined above through numerical simulations.

For each considered vaccine type and bacteria, we computed the relative reduction in the cumulative infection incidence over one year, as well as the total number of averted infections per 100 000 individuals over one year, and the relative change in resistance proportion, as compared with the non-vaccination scenario. These analyses were performed considering vaccine coverages ranging from 10 to 90% and for vaccine efficacies of 30, 60 and 90%.

Moreover, we computed the basic reproduction number *R*_0_ and identified, for each type of vaccine and each vaccine efficacy, the minimum vaccine coverage needed to reach *R*_0_ *<* 1 (i.e. the vaccination threshold).

Finally, we compared the impact of vaccination to that of a gold-standard intervention against resistance, that is, antibiotic stewardship which would translate to a 10 to 90% reduction in bystander antibiotic exposure.

All analyses were performed in R 4.4.2. For numerical simulations, we used the lsoda method of the function ode from the package desolve. PRCC analyses were done using the R packages lhs and epiR. For parallel computations, we used the R packages furrr and parallel. For visualisation, we used ggplot2, ggh4x, scales and patchwork. All codes for this study are available at https://github.com/chloeaupepin/generic_model_paper.

## 3 Results

### 3.1 Conditions for coexistence

All assumptions mentioned in this section are detailed in Table 2, and detailed computations are provided in Appendix A.2.

In the absence of vaccination and of antibiotics (Assumption 1), or of antibiotic-induced resistance selection (Assumption 2), coexistence between the sensitive and resistant strains happens only for a unique value of the relative fitness, respectively:

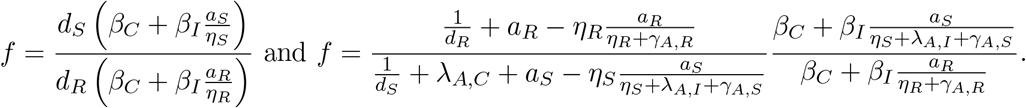

When the entire population is vaccinated and no antibiotics are used (Assumption 4), or antibiotics are used but only provide a decolonising effect without selecting for resistance (Assumption 5), the previous conditions for coexistence are modulated and become:

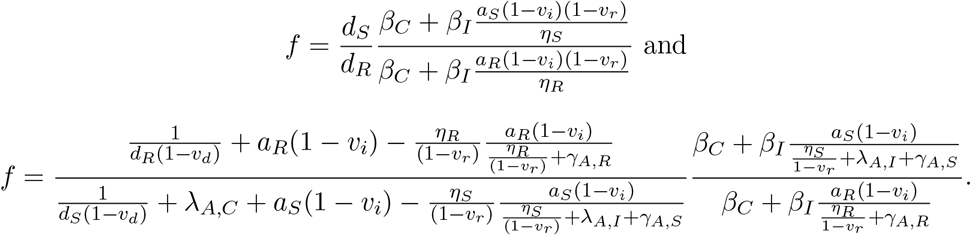

Thus under these conditions, coexistence only occurs if the relative fitness exactly counterbalances the penalty on sensitive strains due to clearance via antibiotic exposure.

For Assumption 3, where antibiotics can select for resistant bacteria but there is no vaccination, we found that coexistence can happen if two conditions are met, which we interpreted in terms of the basic reproduction numbers of sensitive (*R*_0*S*_) and resistant bacteria (*R*_0*R*_) in the absence of the other strain (see Appendix A.1 for detailed computations of these reproduction numbers). The two conditions are

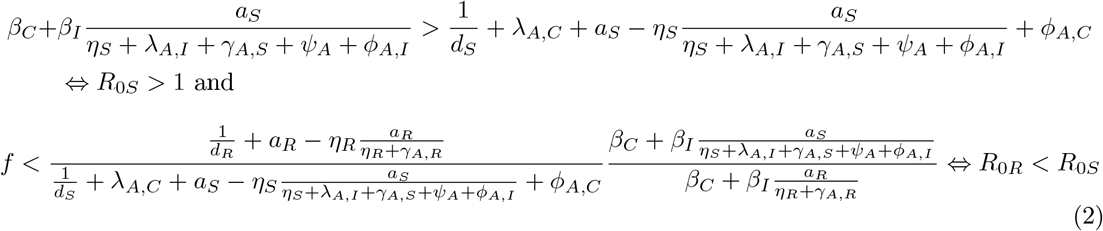

The first condition corresponds to the ability of bacteria to survive (i.e. they must spread faster than they disappear). The second can be interpreted as a threshold for the relative fitness of resistant bacteria, above which they take over and the sensitive strain is eradicated. We also note that the second condition is a relaxation of Assumption 2, whereby there will be coexistence for all values of *f* underneath a threshold.

Finally, when a vaccine is introduced, and the entire population is vaccinated (Assumption 6), these conditions become

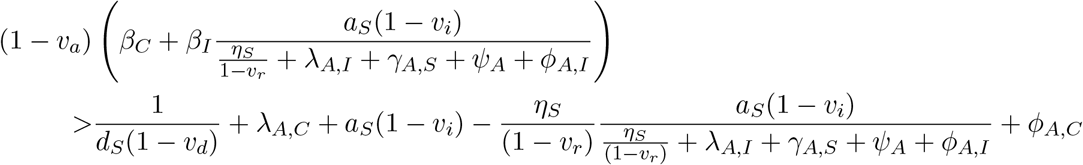

and

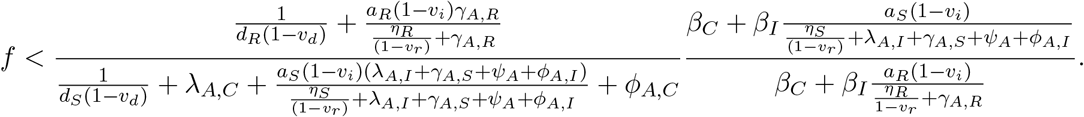

Crucially, the vaccine modifies the conditions, which means that if initially two strains were coexisting, the introduction of a vaccine could derail this coexistence equilibrium. Depending on the parameter values, a vaccine that acts identically on the resistant and sensitive strain can either maintain coexistence at different levels, eliminate the sensitive strain and maintain only the resistant strain, or eliminate both strains altogether. However, we note that a vaccine equally impacting the two strains cannot eliminate only the resistant strain.

To summarise, we found that coexistence equilibrium can only be reliably achieved if antibiotic exposure with a selection rate for resistant bacteria is included in the model. With this selection process, some individuals colonised by a sensitive strain and a minority resistant strain become predominantly colonised by a resistant strain following selection of this minority strain due to antibiotic exposure.

### 3.2 Key parameters

The key parameters influencing the predicted total cumulative incidence of infections are the transmission rate from colonised individuals *β*_*C*_, colonisation duration *d* and infection rate *a* (first column of Figure 2).

**Figure 2:**
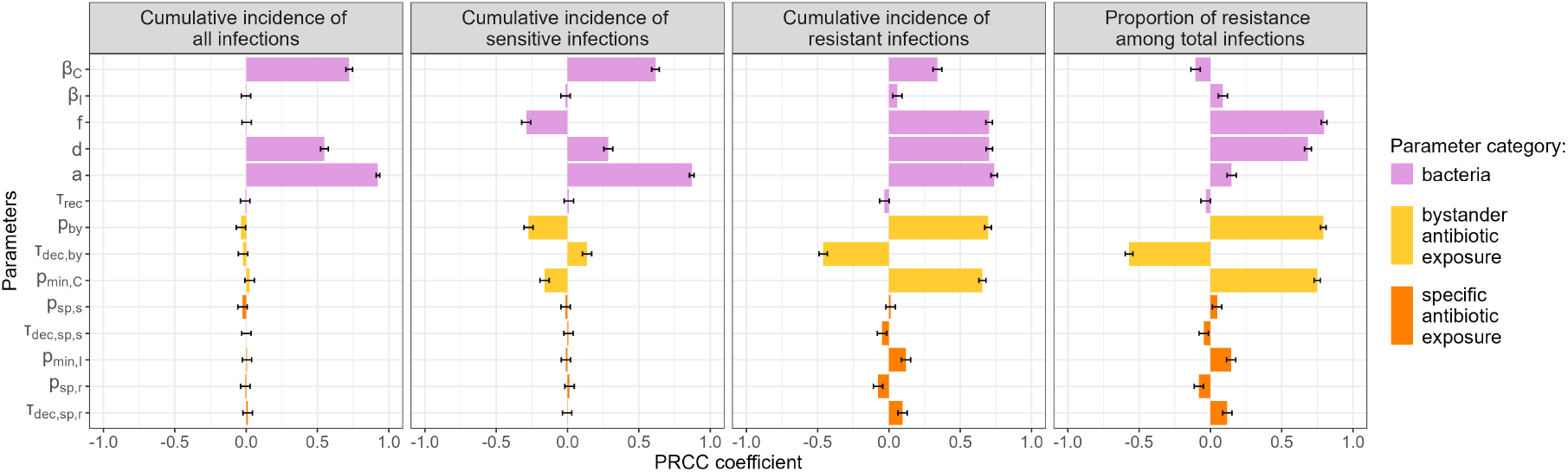
Partial rank correlation coefficients (PRCC) for different outputs of a simulation until equilibrium without vaccine. Parameter ranges considered are presented in Table A1 and explored through Latin Hypercube Sampling (5 000 samples). PRCC indicates the monotony between a specific parameter and an output variable. Positive values correspond to an increase, negative values to a decrease.

Five parameters influence the relative equilibrium between sensitive and resistant strain (last three columns of Figure 2). Resistant strains benefit from an increase of the relative fitness *f*, the rate of bystander antibiotic exposure *p*_*by*_, the probability of minority resistant strain selection *p*_*min,C*_ and the colonisation duration *d*. All four have a positive partial rank correlation coefficient (PRCC) between 0.6 and 0.75 for the cumulative incidence of resistant infections (and the associated proportion of resistance among total infections) and the first three have a negative PRCC for sensitive infections. On the contrary, the duration before antibiotic-induced bacterial clearance *τ*_*dec,by*_ advantages sensitive strains (positive PRCC for sensitive infections, negative for resistant ones).

The vaccine parameters which substantially affect the overall vaccine impact on infections (total, sensitive, resistant) are the percentage of vaccinated individuals (*V*_*perc*_), the reduction of acquisition (*v*_*a*_), the acceleration of the decolonisation (*v*_*d*_) and the reduction of the infection rate (*v*_*i*_) (Figure 3). Increasing these parameters increases the relative reduction of all cumulative incidences. We also observe a negative PRCC associated to the reduction of acquisition (*v*_*a*_) for the relative change in resistance proportions (last column of Figure 3), which means that the vaccine is less efficient to control resistance when it reduces transmission to a greater extent.

**Figure 3:**
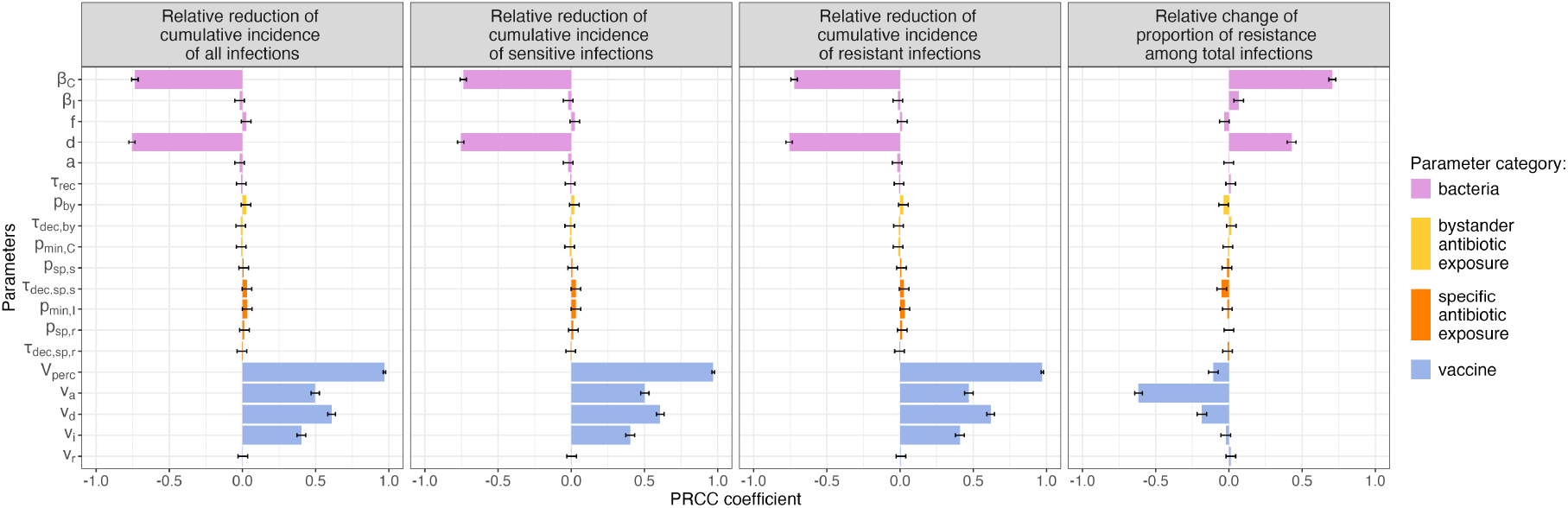
Partial rank correlation coefficients (PRCC) for different outputs of the comparison between one year simulations without vaccine and with vaccine. Parameter ranges considered are presented in Table A1 and explored through Latin Hypercube Sampling (5 000 samples). PRCC indicate the monotony between a specific parameter and an output variable. Positive values correspond to an increase, negative values to a decrease.

The only two bacterial parameters which affect the impact of the vaccine are the transmission rate *β*_*C*_ and the colonisation duration *d* (pink bars on Figure 3). An increase in these bacterial parameters translates to an increase in post-vaccination resistance proportions, and a decrease in all other outputs.

Similar effects of bacterial and vaccine parameters are observed when considering the prevalence of colonisation (total, sensitive, resistant) instead of infection, with the exception of the infection rate *a* and the effect of the vaccine on this infection rate *v*_*i*_, which, as expected, have no influence on colonisation-based outcomes (Figures A2 and A3).

### 3.3 Exploration of vaccine impacts

#### 3.3.1 Vaccines reduce infections

##### In the entire population

As noted in Figure 3, the possible vaccine effect on the recovery rate (*v*_*r*_) does not influence our outcomes of interest. We therefore excluded this vaccine effect for subsequent analysis, ending up with 7 different vaccine types to compare (V_a_, V_d_, V_i_, V_a,d_, V_a,i_, V_d,i_ and V_a,d,i_). Figure 4 depicts the predicted impact of these vaccines on the annual cumulative incidence of total infections for the two bacterial case studies we considered. The impact on the annual cumulative incidence of resistant infections is presented in Figure A4. For both bacteria, all vaccines reduce these cumulative incidences in the total population.

**Figure 4:**
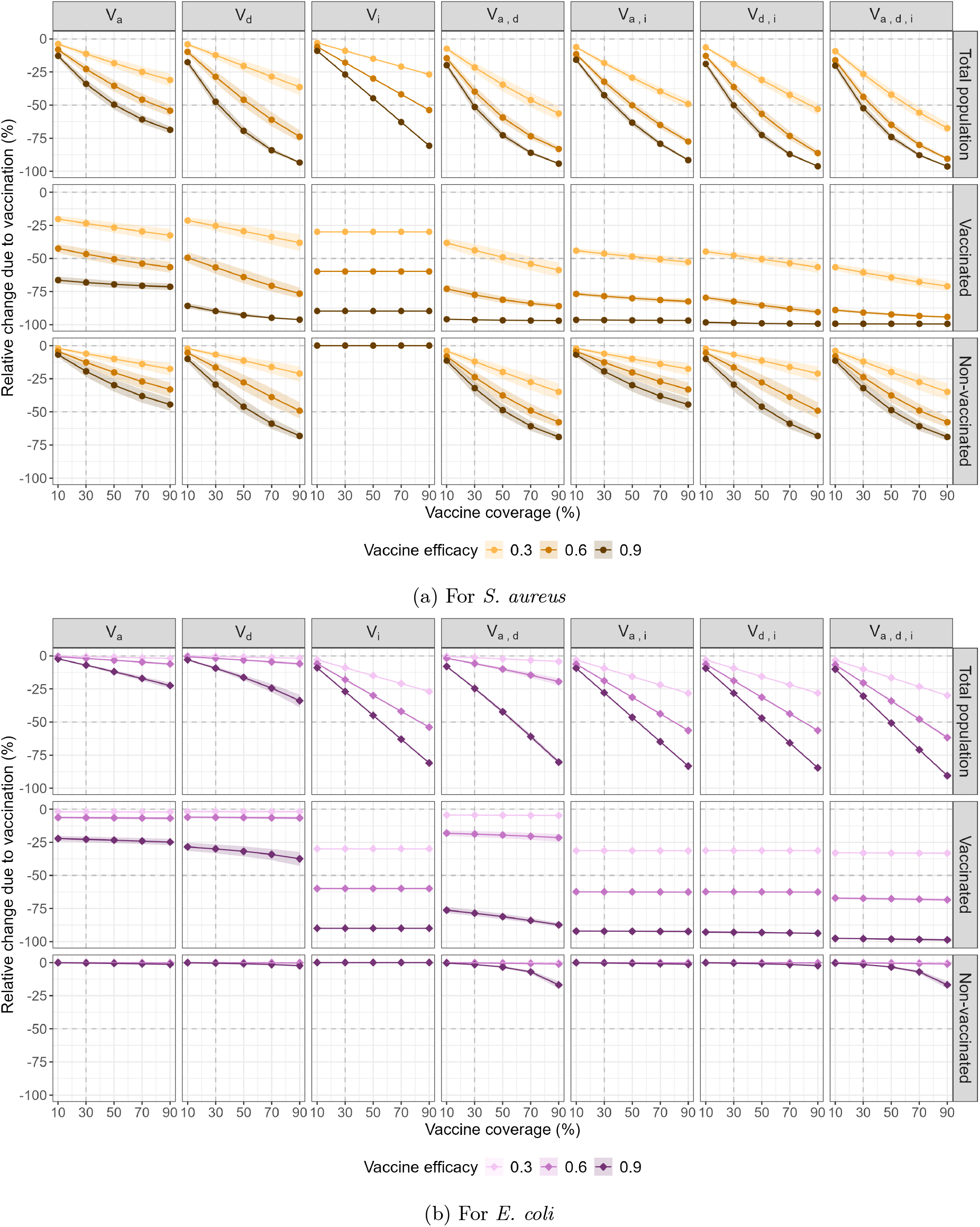
Impact of vaccines on the annual cumulative incidence of total infections for two bacteria. The points indicate the median relative reduction in the predicted incidence as a function of vaccine coverage, as compared with the no-vaccine incidence, and the ribbon depicts the 95% interval between the 2.5th and 97.5th percentiles. Each column corresponds to one vaccine type defined in Table 1. For each considered vaccine, three levels of efficacy are considered: 30% (light), 60% (medium) and 90% (dark), assuming identical vaccine-driven impacts on model parameters for vaccines that incorporate several. Each row corresponds to a specific population (total, vaccinated or non-vaccinated).

On Figure 4a representing the vaccine-induced relative change in bacteraemia incidence caused by *S. aureus*, the best vaccine is the one combining all 3 effects. Considering an arbitrary objective to reduce all infection incidence in the total population (first row) by 50%, for a vaccine efficacy value of 0.3 (yellow lines), the vaccines V_a_,V_d_, V_i_ and V_a,i_ never reach this objective even at 90% coverage, the vaccines V_a,d_ and V_d,i_ need between 70% and 90% coverage to reach this objective, whereas the V_a,d,i_ vaccine only needs a coverage of approximately 60% to reach this target.

The qualitative impact of vaccination is similar for UTIs caused by *E. coli* (Figure 4b), but the reduction in the cumulative incidence of infections is lower for *E. coli* than for *S. aureus*. In terms of absolute numbers of infections however, vaccine impacts are much larger in *E. coli*, with predicted numbers of infections averted up to 100 times higher for *E. coli* UTIs than for *S. aureus* bacteraemia (see Figure 5). This is explained by the much higher baseline incidence of UTIs in the population.

**Figure 5:**
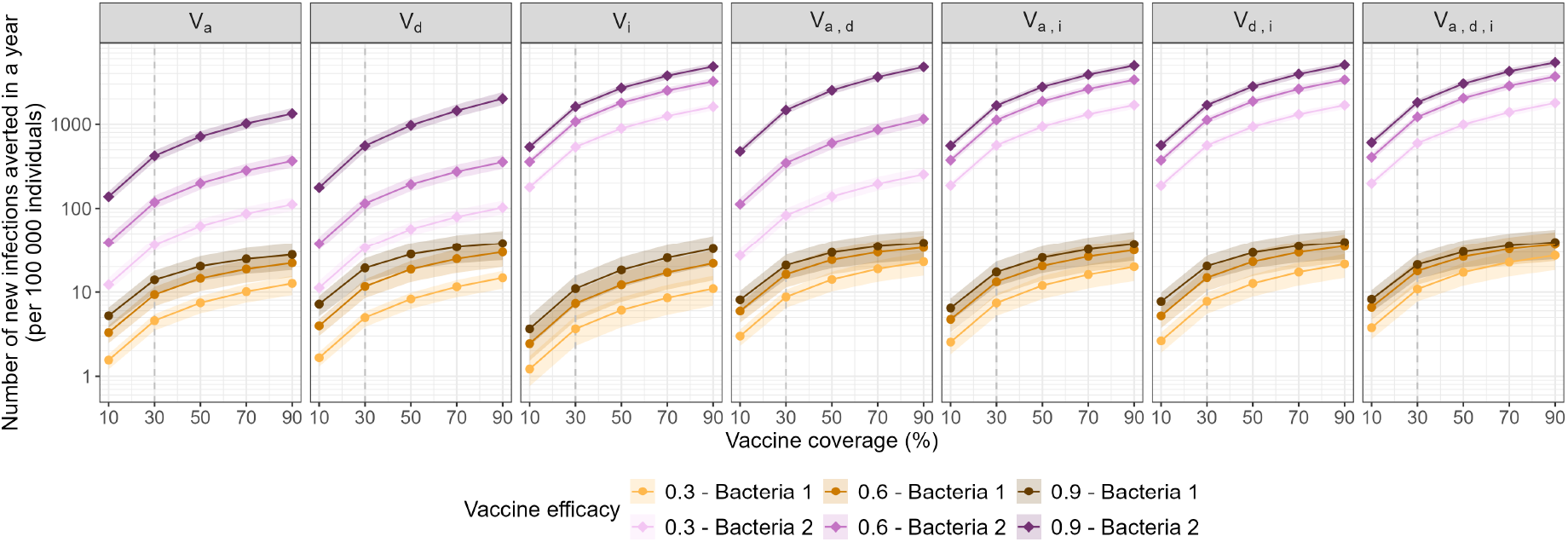
Number of new infections averted over one year by the vaccine in the total population (per 100 000 individuals). Bacteria 1 refer to *S. aureus* and bacteria 2 to *E. coli*. The points indicate the median number of averted infections by a vaccine as a function of vaccine coverage, the ribbon depicts the 95% interval between the 2.5th and 97.5th percentiles. Each column corresponds to one vaccine type defined in Table 1. For each considered vaccine, three levels of efficacy are considered: 30% (light), 60% (medium) and 90% (dark), assuming identical vaccine-driven impacts on model parameters for vaccines that incorporate several.

##### In sub-populations

Focusing on the vaccinated population only, all vaccines reduce the cumulative infection incidence (Figure 4a, second row). In that sub-population, a reduction of 50% of the cumulative incidence is easier to reach and most vaccines (except for V_a_, V_d_, and V_i_) reach this objective with at least 50% coverage and 30% efficacy. Lower relative reductions are again observed for *E. coli* (Figure 4b, second row).

The non-vaccinated sub-population may be affected through herd immunity if coverage is sufficiently high [35]. We observe differing patterns between *S. aureus* and *E. coli*. All considered vaccines reduce the cumulative incidence of *S. aureus* bacteraemia in the non-vaccinated sub-population, except, as expected, for the vaccine which only reduces infection rates (V_i_) and has no impact on colonisation (Figure 4a, third row). For *E. coli* UTIs however, the indirect effect on non-vaccinated individuals is practically inexistent for all vaccine types considered (Figure 4b, third row).

#### 3.3.2 Vaccines may reduce colonisation prevalence

Vaccines reducing acquisition or accelerating natural clearance reduce colonisation prevalence (Figure A5). As expected, vaccines reducing only the risk of infection do not impact colonisation prevalence.

Figure A6 presents, for each bacterium and each vaccine type, the basic reproduction number (*R*_0_). For *S. aureus, R*_0_ decreases linearly as vaccine coverage increases, for all vaccine types except V_i_. For each vaccine type and for a given vaccine efficacy, we can derive the minimum coverage required to bring *R*_0_ below 1, and therefore control the spread of the bacterium. For example, for a vaccine reducing acquisition (V_a_) with 60% efficacy, a 49% coverage across the entire population is sufficient to control the spread of the bacteria (Figure A6).

For *E. coli*, the vaccine V_i_ increases *R*_0_ while all other vaccine types decrease it. Controlling its spread is more difficult than for *S. aureus*, since only a vaccine which both decreases acquisition and increases natural clearance with 90% efficacy and 96% coverage can bring the *R*_0_ below 1.

#### 3.3.3 Vaccines may increase or decrease the proportion of resistant carriers

Under certain conditions, vaccines can increase the proportion of resistant colonisation among all colonised. For *S. aureus*, this occurs with all vaccines except the one that exclusively reduces the infection rate as observed on Figure 6a. This increase in the proportion of resistant colonised is observed among both vaccinated and non-vaccinated individuals. In the total population, the increase can reach up to 26% for the vaccines V_a,d_ and V_a,d,i_. However, at low efficacy levels, vaccines V_d_ and V_d,i_ slightly decrease the resistant proportion instead of increasing it. In contrast, when applied to *E. coli*, the same vaccines can reduce the proportion of resistant colonised in the total population by up to 44% for V_d_ and V_d,i_ (Figure 6b).

**Figure 6:**
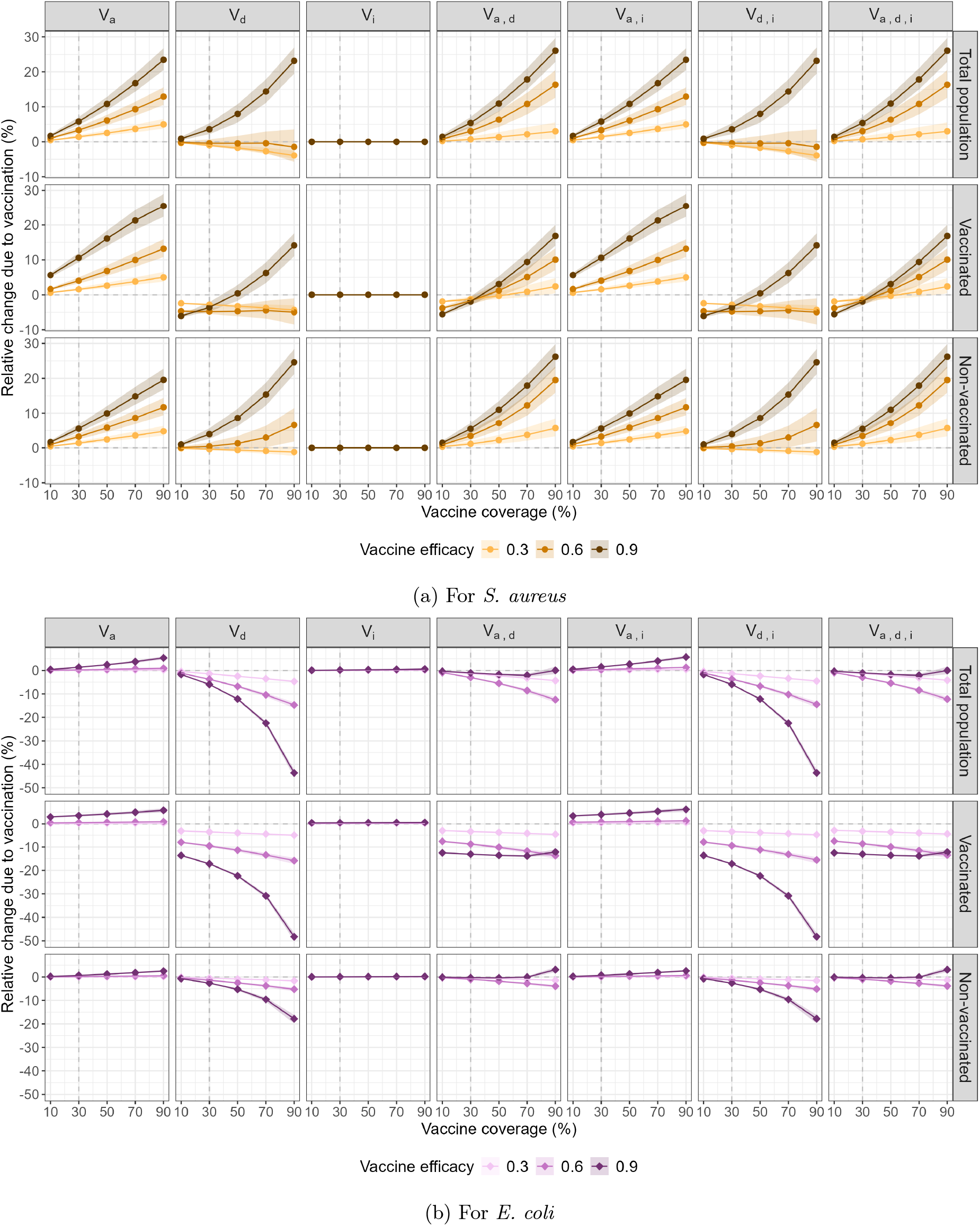
Impact of vaccines on the proportion of resistant colonisation among colonised. The points indicate the median relative reduction in the predicted proportion of resistance as a function of vaccine coverage, as compared with the no-vaccine proportion, and the ribbon depicts the 95% interval between the 2.5th and 97.5th percentiles. Each column corresponds to one vaccine type defined in Table 1. For each considered vaccine, three levels of efficacy are considered: 30% (light), 60% (medium) and 90% (dark), assuming identical vaccine-driven impacts on model parameters for vaccines that incorporate several. Each row corresponds to a specific population (total, vaccinated or non-vaccinated).

### 3.4 Comparison with bystander antibiotic exposure reduction

Bystander exposure reductions always decreased both the proportion of resistant infections and the proportion of resistant colonised, an outcome which was not consistently observed with vaccines (Figure 7, last column). For *S. aureus*, a 90% reduction in bystander exposure led to a 23% reduction in the proportion of resistant colonised, outperforming all vaccine types considered. For *E. coli* this same reduction in bystander exposure led to a 12% reduction of the proportion of resistant colonised, a reduction more than 2-fold inferior to that obtainable with highly effective vaccines accelerating decolonisation.

**Figure 7:**
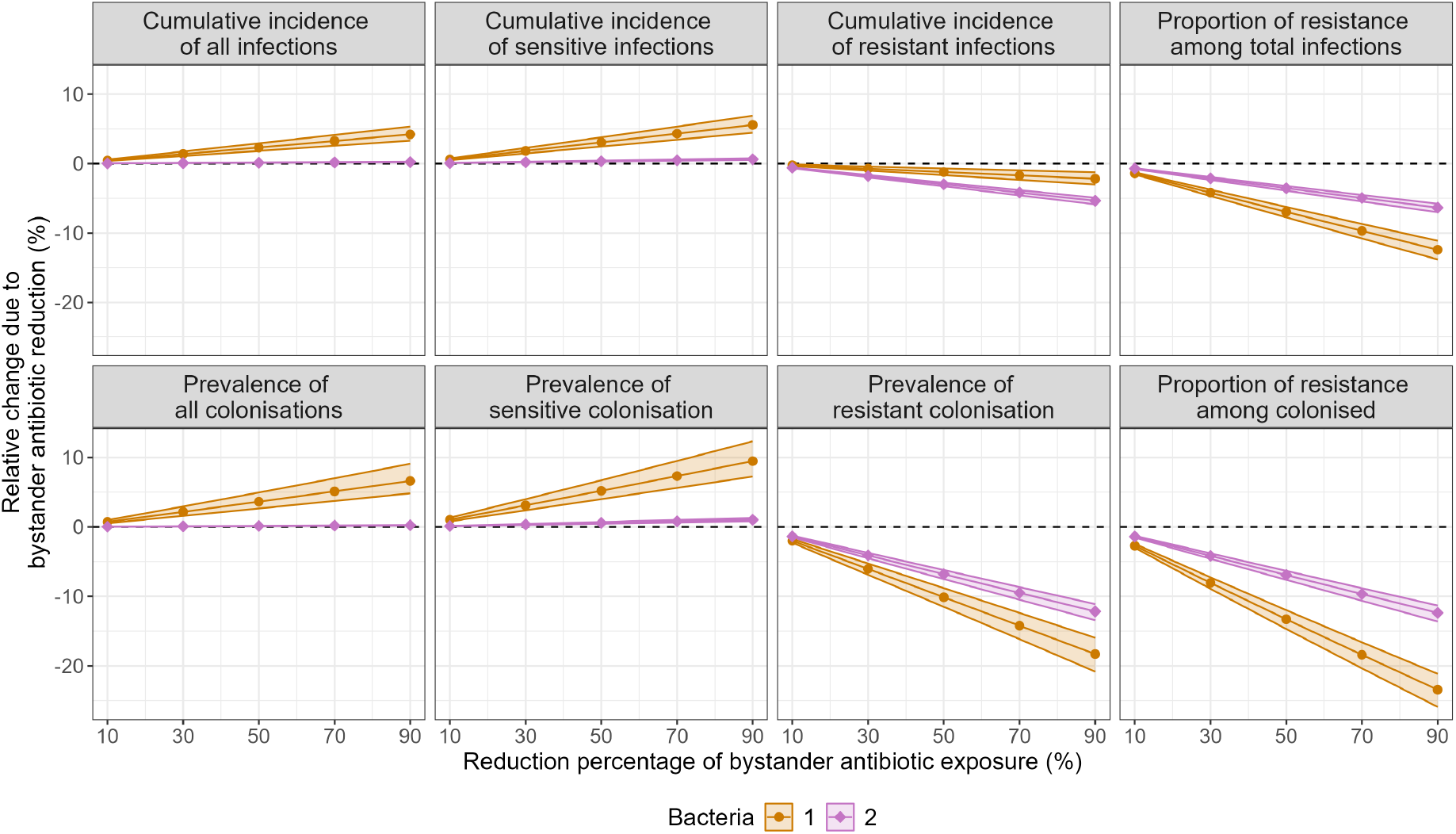
Impact of bystander exposure reduction for different outputs on the total population. Bacteria 1 refer to *S. aureus* and bacteria 2 to *E. coli*. The points indicate the median relative reduction in the predicted incidences and prevalences as a function of bystander exposure reduction, as compared with the baseline bystander exposure, and the ribbon depicts the 95% interval between the 2.5th and 97.5th percentiles.

Moreover, while vaccines consistently reduced the incidence of infections, reductions in bystander antibiotic exposure comparatively showed an increase on overall incidence of infections and prevalence of colonisation for *S. aureus* and little effect for *E. coli* (Figure 7, first column). Effectively, the increase in sensitive infections and colonisations due to the reduced bystander selective pressure exactly counteracted the decrease in resistant infections and colonisations for *E. coli* but was too strong for *S. aureus* (Figure 7, second and third column).

## 4 Discussion

In this study, we proposed a novel generic mathematical framework to explore the potential impact of bacterial vaccines on antibiotic resistance. Our analytical results showed that vaccines can change the conditions under which an equilibrium with coexistence of sensitive and resistant strains exists. Moreover, based on numerical simulations of vaccine introduction, we observed that, while vaccines may reduce the absolute number of resistant infections and sensitive infections, they may also increase the resistance proportion among colonised in the population. These impacts were shown to depend strongly on the characteristics of the considered bacteria, resistance and infection.

Bacteria of public health interest typically coexist as sensitive and resistant strains at the population level [19, 20, 21]. We showed that this equilibrium can only be achieved if antibiotic use selects for a minority resistant strain. This feature is essential to maintain a balance between resistant and sensitive bacteria, since antibiotic use disadvantages sensitive strains, whereas resistant strains are disadvantaged by their lower relative fitness. Selection for a minority resistant strain further balances these effects, turning the disadvantage of sensitive strains under antibiotic pressure into an advantage for resistant strains and thereby allowing for stable coexistence over a wide range of plausible conditions. Spicknall and colleagues found an equivalent result with models that did not differentiate between colonisation and infection. Their *unidirectional conversion* model - corresponding to what we refer to as antibiotic selection - allowed for coexistence when the reproductive number of the resistant strain was lower than that of the sensitive strain, which aligns with our condition (2) [36].

For both *S. aureus* and *E. coli*, we showed that vaccines have the potential to reduce the number of resistant infections (Figure A4), as well as the overall number of infections (Figure 4). This motivates further research on these vaccines. In the earlier analysis by the WHO, a vaccine against *S. aureus* targeting all people with an efficacy of 60% and a coverage of 70% was estimated to reduce deaths associated with resistance in 2019 by 40% in Europe [18]. Using our model to replicate this scenario, with only an efficacy to reduce infection risk by 60% and a coverage of 70%, we find that the cumulative incidence of resistant bacteraemia is reduced by 41%. Assuming that the number of deaths associated to resistance in *S. aureus* is linearly linked to the number of severe resistant infections such as bacteraemia, our estimation therefore aligns with WHO results. However, our results also suggest that a vaccine combining the three effects we considered (on acquisition, decolonisation, and infection), with each corresponding vaccine efficacy parameter fixed at 60% and a vaccine coverage of 70%, could reduce the cumulative incidence of resistant bacteraemia by 78%, a substantial increase compared to the WHO results (Figure A4a). This is explained by the fact that our model includes mechanisms of transmission and herd immunity that could not be taken into account by a static model such as the one used by WHO. For *E. coli*, the WHO found that a 70% coverage of a vaccine with a 70% efficacy against urinary tract infections could reduce deaths associated with resistance in 2019 by approximately 50%. Again, by replicating this scenario using our model we obtain a similar results (49% reduction), but allowing for 70% efficacy for all three vaccine effects yields a 58% incidence reduction instead (Figure A4b). Our work therefore highlights the importance of taking indirect effects into account when quantifying vaccine impacts on ABR.

In terms of public health, it is useful to know in advance what vaccine coverage is needed to control the spread of a disease for a specific vaccine. One novelty of our proposed framework lies in the ability to estimate this vaccine coverage while accounting for a range of possible vaccine effects. For instance, with a vaccine acting on acquisition and infection with a 60% efficacy, a 49% vaccine coverage across the entire population suffices to control the spread of *S. aureus* (Figure A6a). On the other hand, due to a high colonisation prevalence, *E. coli* transmission could only be controlled by a vaccine combining at least both effects on acquisition and decolonisation with a 96% vaccine coverage. In this situation and without further interventions, *E. coli* colonisation could decline and reach a 0% prevalence. In practice, this outcome is likely not desirable due to the usefulness of these commensal bacteria within the gut microbiota [37]. However, if another high-colonising bacteria were responsible for a substantial public health burden without being beneficial, it is important to recognise that achieving epidemic control would require really high vaccination coverage.

We compared the impact of the introduction of a vaccine to that of a reduction in by-stander antibiotic reduction since this corresponds to a scenario of antibiotic stewardship, a key intervention against antibiotic resistance [38]. We found that antibiotic reduction always lowered the proportion of resistant infections and colonisations, but could increase the overall colonisation prevalence by giving an advantage to the sensitive strain and could thus increase the number of infections (Figure 7). In comparison, vaccines could reduce overall colonisations and infections (Figures A5 and 4) but could increase the proportion of resistance (Figure 6). This highlights the necessity to define precise criteria to identify an optimal public health intervention strategy. If the goal is to reduce the resistance proportion, bystander antibiotic reduction and vaccines acting only on decolonisation are more appropriate, but if the need is to reduce all infections, vaccines appear to be the best choice, regardless of their mechanisms of action.

By focusing on two specific case studies, we showed how identical vaccines can have very different impacts depending on the bacteria studied. As seen in Figure 6, vaccines accelerating decolonisation tend to reduce the resistance proportion among colonised for *E. coli* while increasing it for *S. aureus*. This contrast may be explained by differences in colonisation duration, which was fixed at 365 days for *E. coli* and 98 days for *S. aureus*. Longer colonization durations provide more time for bystander selection to occur; therefore, reducing colonisation duration is expected to reduce bystander selection pressure, which could account for the decrease observed in the resistance proportion among colonised for *E. coli*. On the other hand, the increase observed for *S. aureus* may be explained by the reduction in prevalence of sensitive colonised by the vaccine, leading to more opportunities for resistant bacteria acquisition. Likewise, the difference observed when reducing antibiotic bystander exposure may be due to a difference in the magnitude of bystander exposure being almost 3 times higher for *S. aureus*. This highlights the utility of our generic model, which is able to capture how intervention impacts may vary across different case studies.

Although our analysis focused on two specific case studies, the structure of the model is intentionally designed to be broadly applicable across a wide range of bacterial pathogens, associated infections, resistances and potential vaccines. This is particularly relevant in the context of uncertainty regarding epidemiological data on bacterial infections and the resulting model parameter values. As we have shown through our sensitivity analyses, our model can explore a wide range of values to provide guidance in highly uncertain context on which parameters are likely to have a greater impact on bacterial dynamics. Our model takes into account colonised and infected states as compartments. Thus, it can be applied to bacteria with dynamics either governed by colonised or infected individuals, or both. Moreover, taking into account both states enables us to consider both bystander antibiotic exposure and specific antibiotic exposure which, to our current knowledge, has not been done before [10]. The model further integrates both selection for resistant strains and clearance of sensitive strains by antibiotics, enabling a more mechanistic representation of treatment effects on population-level resistance dynamics. With respect to vaccination, we designed a flexible structure including four possible distinct vaccine effects that allows a differentiation between sensitive or resistant bacteria. Multi-functionality is particularly relevant to test different hypotheses and improve the systematic exploration of vaccine effects on antibiotic resistance, since different vaccines may impact different aspects of bacterial dynamics. This makes our model adaptable to any future vaccine implementation and thus increases its usefulness to determine overall impact of bacterial vaccines on antibiotic resistance.

Nonetheless, our work has certain limitations that should be taken into account.

The model was designed to represent a large variety of bacterial dynamics, which entailed some necessary simplifications. Co-colonisation was integrated in a minimal form thanks to the possibility for an antibiotic to select for a minority resistant strain. However, in some instances, co-colonisation may be more extensive and complex, as seen for *S. agalactiae* [39]. Some other mathematical models include additional compartments to represent the simultaneous presence and interaction of multiple strains [40]. Furthermore, for bacteria with multiple cocirculating serotypes such as *S. pneumoniae*, vaccination may lead to serotype replacement. This phenomenon can be reproduced by including compartments for both vaccine and non-vaccine serotypes in the model, which was not considered here for the sake of generality [41].

We considered only two discrete states for bacterial strains: either sensitive or resistant to antibiotics. In this simplified framework, antibiotics were assumed to be completely effective against sensitive strains and entirely ineffective against resistant strains. However, the nature of resistance depends on the underlying mechanism. Some resistance mechanisms indeed produce an essentially binary (on/off) phenotype, while others confer partial resistance, leading to a continuum of susceptibilities. For this latter case, some modelling studies propose to represent resistance on a gradual scale, although this again comes at the cost of a higher number of compartments and therefore difficulties to analytically study the model [11, 13].

The compartmental structure of our model is deterministic and represents average dynamics, making it well-suited to community-level transmission. The structure could be adapted to bacteria that develop predominantly within healthcare environments. This would involve taking into account individual variabilities (such as co-morbidities for patients, or job for healthcare workers) and the stochasticity of contacts in hospitals [14]. The model currently assumes a closed population with homogeneous mixing well-suited to study the entire population. It could also be applied to a specific subpopulation provided that the transmission occurs exclusively within that subpopulation.

Another possible adaptation of the model would be age stratification. Differences in ABR prevalence by ages have been observed in Europe for bloodstream infections [42], suggesting that transmission and selection dynamics vary with age. Incorporating an age structure into the model would lead to a more accurate representation of these dynamics, as well as the evaluation of targeted vaccination strategies on certain subpopulations and a more precise understanding of the impact of vaccines on ABR according to age. However, this improvement would need to be informed with more precise data to obtain age-specific transmission rates, risks of infection and antibiotic prescriptions.

Finally, we currently assume constant immunity and immediate vaccine efficacy. While valid for one-year simulations, these assumptions would need adjustment for longer-term scenarios. Accounting for waning immunity and dynamic vaccination strategies would allow us to evaluate more precisely the population-level impact of vaccines. This would allow us to anticipate temporal variations of vaccine impact on ABR thus translating our results to more precise public health recommendations.

In summary, our study offers a theoretical understanding of how bacterial vaccines might affect antibiotic resistance while taking into consideration bacterial transmission and antibiotic selection dynamics. By providing quantitative insights about the expected effects of various vaccines as well as a generic, easily adaptable framework to study a diverse range of bacteria and vaccines, our study enables us to compare intervention strategies, thereby facilitating public health decision-making and contributing to the development of an effective response to antibiotic resistance.

## Data Availability

All data produced in the present work are contained in the manuscript.

## Data Accessibility

This article has no additional data.

## Authors’ Contributions

CA: formal analysis, investigation, methodology, software, validation, visualization, writing – original draft, writing – review & editing; LO: conceptualization, investigation, methodology, validation, writing – review & editing; IvB: data curation, writing – review & editing; ES: data curation, writing – review & editing; VS: data curation, writing – review & editing; SL: formal analysis, investigation, methodology, supervision, validation, visualization, writing – original draft, writing – review & editing; LT: conceptualization, formal analysis, investigation, methodology, supervision, validation, visualization, writing – original draft, writing – review & editing; QL: conceptualization, formal analysis, investigation, methodology, supervision, validation, visualization, writing – original draft, writing – review & editing

