## Supplementary material for "Quantifying the impact of bacterial vaccines against antibiotic resistance: accounting for transmission and selection dynamics"

**References** **21**

### A Appendix

#### A.1 Reproduction number computation

The basic reproduction number, denoted $R_0$, is a key epidemiological metric used to describe the contagiousness of an infectious disease. It represents the average number of secondary infections produced by one infected individual in a completely susceptible population. We used the method explained by O. Diekmann and colleagues [1] and P. van den Driessche and J. Watmough [2]. $R_0$ is the spectral radius of the next generation matrix denoted $G$. This matrix is the product of the transmission matrix $F$ and the inverse of the transition matrix $V$. The transmission matrix and the transition matrix correspond respectively to the partial derivatives of the rates of new colonisations ($\mathcal{F}$) and of the rates of change in infected compartments ($\mathcal{V}$) with respect to the infected states (specifically ordered $C_S^{nv}, C_S^v, I_S^{nv}, I_S^v, C_R^{nv}, C_R^v, I_R^{nv}, I_R^v$) evaluated at the disease-free equilibrium.

The disease-free equilibrium is given by

$$C_S^{nv*} = C_R^{nv*} = I_S^{nv*} = I_R^{nv*} = C_S^{v*} = C_R^{v*} = I_S^{v*} = I_R^{v*} = 0,$$

$U^{nv*} = N^{nv}$ and $U^{v*} = N^v$, where $N^{nv}$ is the total number of non-vaccinated individuals and $N^v$ the total number of vaccinated individuals. We denote $V_{perc}$ the vaccine coverage so $N^{nv} = (1 - V_{perc})N$ and $N^v = V_{perc}N$ where $N$ is the total population size.

We now separate model equations governing infected states into transmission effects and transition effects:

$$\begin{cases}
\frac{dC_S^{nv}}{dt} &= B_S U^{nv} - \left(\frac{1}{d_S} C_S^{nv} + \lambda_{A,C} C_S^{nv} + \phi_{A,C} C_S^{nv} + a_S C_S^{nv} - \eta_S I_S^{nv}\right) \\
&= \mathcal{F}_1 + \mathcal{V}_1 \\
\frac{dC_R^{nv}}{dt} &= B_R U^{nv} - \left(\frac{1}{d_R} C_R^{nv} - \phi_{A,C} C_S^{nv} + a_R C_R^{nv} - \eta_R I_R^{nv}\right) \\
&= \mathcal{F}_5 + \mathcal{V}_5 \\
\frac{dI_S^{nv}}{dt} &= -(-a_S C_S^{nv} + \eta_S I_S^{nv} + \lambda_{A,I} I_S^{nv} + \gamma_{A,S} I_S^{nv} + \psi_A I_S^{nv} + \phi_{A,I} I_S^{nv}) \\
&= \mathcal{F}_3 + \mathcal{V}_3 \\
\frac{dI_R^{nv}}{dt} &= -(-a_R C_R^{nv} + \eta_R I_R^{nv} - \psi_A I_S^{nv} - \phi_{A,I} I_S^{nv} + \gamma_{A,R} I_R^{nv}) \\
&= \mathcal{F}_7 + \mathcal{V}_7 \\
\frac{dC_S^v}{dt} &= B_S^v U^v - \left(\frac{1}{d_S(1-v_d)} C_S^v + \lambda_{A,C} C_S^v + \phi_{A,C} C_S^v + a_S(1-v_i) C_S^v - \frac{\eta_S}{1-v_r} I_S^v\right) \\
&= \mathcal{F}_2 + \mathcal{V}_2 \\
\frac{dC_R^v}{dt} &= B_R^v U^v - \left(\frac{1}{d_R(1-v_d)} C_R^v - \phi_{A,C} C_S^v + a_R(1-v_i) C_R^v - \frac{\eta_R}{1-v_r} I_R^v\right) \\
&= \mathcal{F}_6 + \mathcal{V}_6 \\
\frac{dI_S^v}{dt} &= -\left(-a_S(1-v_i) C_S^v + \frac{\eta_S}{1-v_r} I_S^v + \lambda_{A,I} I_S^v + \gamma_{A,S} I_S^v + \psi_A I_S^v + \phi_{A,I} I_S^v\right) \\
&= \mathcal{F}_4 + \mathcal{V}_4 \\
\frac{dI_R^v}{dt} &= -\left(-a_R(1-v_i) C_R^v + \frac{\eta_R}{1-v_r} I_R^v - \psi_A I_S^v - \phi_{A,I} I_S^v + \gamma_{A,R} I_R^v\right) \\
&= \mathcal{F}_8 + \mathcal{V}_8
\end{cases}$$

This constructs the vector $\mathcal{F}$ of the rates of new colonisations and the vector $\mathcal{V}$ of transitions between colonised and infected compartments. For better readability, equilibrium values of compartments are denoted without the star until further notice. We also introduce $\lambda_X = \frac{1}{d_X}$ the inverse of colonisation duration , $\lambda_X^v = \frac{\lambda_X}{1-v_d}$ the vaccine-modified decolonisation rate, $a_X^v = a_X(1 - v_i)$ the vaccine-modifier infection rate and $\eta_X^v = \frac{\eta_X}{1-v_r}$ the vaccine-modified recovery rate (where $X = S$ or $X = R$ depending on the strain). We denote by $H_{C_S^{nv}}$ the sum of all output flows of the compartment $C_S^{nv}$, thus $H_{C_S^{nv}} = \lambda_S + \lambda_{A,C} + \phi_{A,C} + a_S$. This is generalised for all compartments.

We obtain the matrices

$$F = \begin{pmatrix} F_1 & 0_{(4,4)} \\ 0_{(4,4)} & F_2 \end{pmatrix} \text{ and } V = -\begin{pmatrix} W & 0 \\ -Z & Y \end{pmatrix} \text{ with}$$

$$F_1 = \begin{pmatrix} \beta_C(1-V_{perc}) & \beta_C(1-V_{perc}) & \beta_I(1-V_{perc}) & \beta_I(1-V_{perc}) \\ \beta_C V_{perc}(1-v_a) & \beta_C V_{perc}(1-v_a) & \beta_I V_{perc}(1-v_a) & \beta_I V_{perc}(1-v_a) \\ 0 & 0 & 0 & 0 \\ 0 & 0 & 0 & 0 \end{pmatrix},$$

$$F_2 = \begin{pmatrix} \beta_C f(1-V_{perc}) & \beta_C f(1-V_{perc}) & \beta_I f(1-V_{perc}) & \beta_I f(1-V_{perc}) \\ \beta_C f V_{perc}(1-v_a) & \beta_C f V_{perc}(1-v_a) & \beta_I f V_{perc}(1-v_a) & \beta_I f V_{perc}(1-v_a) \\ 0 & 0 & 0 & 0 \\ 0 & 0 & 0 & 0 \end{pmatrix},$$

$$W = \begin{pmatrix} H_{C_S^{nv}} & 0 & -\eta_S & 0 \\ 0 & H_{C_S^v} & 0 & -\eta_S^v \\ -a_S & 0 & H_{I_S^{nv}} & 0 \\ 0 & -a_S^v & 0 & H_{I_S^v} \end{pmatrix},$$

$$Y = \begin{pmatrix} \lambda_R + a_R & 0 & -\eta_R & 0 \\ 0 & \lambda_R^v + a_R^v & 0 & -\eta_R^v \\ -a_R & 0 & \eta_R + \gamma_{A,R} & 0 \\ 0 & -a_R^v & 0 & \eta_R^v + \gamma_{A,R} \end{pmatrix} \text{ and}$$

$$-Z = \begin{pmatrix} -\phi_{A,C} & 0 & 0 & 0 \\ 0 & -\phi_{A,C} & 0 & 0 \\ 0 & 0 & -\psi_A - \phi_{A,I} & 0 \\ 0 & 0 & 0 & -\psi_A - \phi_{A,I} \end{pmatrix}.$$

We remark that

$$V^{-1} = -\begin{pmatrix} W^{-1} & 0 \\ Y^{-1}ZW^{-1} & Y^{-1} \end{pmatrix} \text{ so } G = -FV^{-1} = \begin{pmatrix} F_1 W^{-1} & 0 \\ F_2 Y^{-1} Z W^{-1} & F_2 Y^{-1} \end{pmatrix}.$$

The matrix $G$ is triangular thus, its eigenvalues are the eigenvalues of the diagonal blocks. We only have to compute $F_1 W^{-1}$ and $F_2 Y^{-1}$.

We obtain

$$W^{-1} = \begin{pmatrix} \frac{H_{I_S^{nv}}}{H_{C_S^{nv}} H_{I_S^{nv}} - a_S \eta_S} & 0 & \frac{\eta_S}{H_{C_S^{nv}} H_{I_S^{nv}} - a_S \eta_S} & 0 \\ 0 & \frac{H_{I_S^v}}{H_{C_S^v} H_{I_S^v} - a_S^v \eta_S^v} & 0 & \frac{\eta_S^v}{H_{C_S^v} H_{I_S^v} - a_S^v \eta_S^v} \\ \frac{a_S}{H_{C_S^{nv}} H_{I_S^{nv}} - a_S \eta_S} & 0 & \frac{H_{C_S^{nv}}}{H_{C_S^{nv}} H_{I_S^{nv}} - a_S \eta_S} & 0 \\ 0 & \frac{a_S^v}{H_{C_S^v} H_{I_S^v} - a_S^v \eta_S^v} & 0 & \frac{H_{C_S^v}}{H_{C_S^v} H_{I_S^v} - a_S^v \eta_S^v} \end{pmatrix} \text{ and}$$

$$Y^{-1} = \begin{pmatrix} \frac{H_{I_R^{nv}}}{H_{C_R^{nv}} H_{I_R^{nv}} - a_R \eta_R} & 0 & \frac{\eta_R}{H_{C_R^{nv}} H_{I_R^{nv}} - a_R \eta_R} & 0 \\ 0 & \frac{H_{I_R^v}}{H_{C_R^v} H_{I_R^v} - a_R^v \eta_R^v} & 0 & \frac{\eta_R^v}{H_{C_R^v} H_{I_R^v} - a_R^v \eta_R^v} \\ \frac{a_R}{H_{C_R^{nv}} H_{I_R^{nv}} - a_R \eta_R} & 0 & \frac{H_{C_R^{nv}}}{H_{C_R^{nv}} H_{I_R^{nv}} - a_R \eta_R} & 0 \\ 0 & \frac{a_R^v}{H_{C_R^v} H_{I_R^v} - a_R^v \eta_R^v} & 0 & \frac{H_{C_R^v}}{H_{C_R^v} H_{I_R^v} - a_R^v \eta_R^v} \end{pmatrix}.$$

We deduce:

$$F_1W^{-1} = \begin{pmatrix} \frac{f_{11}\cdot H_{I_S^{nv}}+f_{13}\cdot a_S}{D_S^{nv}} & \frac{f_{12}\cdot H_{I_S^v}+f_{14}\cdot a_S^v}{D_S^v} & \frac{f_{11}\cdot H_{C_S^{nv}}+f_{13}\cdot \eta_S}{D_S^{nv}} & \frac{f_{12}\cdot H_{C_S^v}+f_{14}\cdot \eta_S^v}{D_S^v} \\ \frac{f_{21}\cdot H_{I_S^{nv}}+f_{23}\cdot a_S}{D_S^{nv}} & \frac{f_{22}\cdot H_{I_S^v}+f_{24}\cdot a_S^v}{D_S^v} & \frac{f_{21}\cdot H_{C_S^{nv}}+f_{23}\cdot \eta_S}{D_S^{nv}} & \frac{f_{22}\cdot H_{C_S^v}+f_{24}\cdot \eta_S^v}{D_S^v} \\ 0 & 0 & 0 & 0 \\ 0 & 0 & 0 & 0 \end{pmatrix} \text{ and}$$

$$F_2Y^{-1} = \begin{pmatrix} \frac{f_{55}\cdot H_{I_R^{nv}}+f_{57}\cdot a_R}{D_R^{nv}} & \frac{f_{56}\cdot H_{I_R^v}+f_{58}\cdot a_R^v}{D_R^v} & \frac{f_{55}\cdot H_{C_R^{nv}}+f_{57}\cdot \eta_R}{D_R^{nv}} & \frac{f_{56}\cdot H_{C_R^v}+f_{58}\cdot \eta_R^v}{D_R^v} \\ \frac{f_{65}\cdot H_{I_R^{nv}}+f_{67}\cdot a_R}{D_R^{nv}} & \frac{f_{66}\cdot H_{I_R^v}+f_{68}\cdot a_R^v}{D_R^v} & \frac{f_{65}\cdot H_{C_R^{nv}}+f_{67}\cdot \eta_R}{D_R^{nv}} & \frac{f_{66}\cdot H_{C_R^v}+f_{68}\cdot \eta_R^v}{D_R^v} \\ 0 & 0 & 0 & 0 \\ 0 & 0 & 0 & 0 \end{pmatrix}$$

where $f_{ij}$ corresponds to the element on ith row and jth column and $D_S^{nv} = H_{C_S^{nv}} H_{I_S^{nv}} - a_S \eta_S$, $D_S^v = H_{C_S^v} H_{I_S^v} - a_S^v \eta_S^v$, $D_R^{nv} = H_{C_R^{nv}} H_{I_R^{nv}} - a_R \eta_R$, and $D_R^v = H_{C_R^v} H_{I_R^v} - a_R^v \eta_R^v$.

We now search the eigenvalues for a matrix of the form

$$\begin{pmatrix} a & b & c & d \\ e & f & g & h \\ 0 & 0 & 0 & 0 \\ 0 & 0 & 0 & 0 \end{pmatrix}.$$

This matrix has the eigenvalue 0 with multiplicity 2 and share eigenvalues with the two by two matrix $\begin{pmatrix} a & b \\ e & f \end{pmatrix}$. The discriminant of the characteristic polynomial of order 2 is:

$$\Delta = (-(a+f))^2 - 4(af - eb)$$

We note that for both $F_1W^{-1}$ and $F_2Y^{-1}$, $af = eb$. We prove it for $F_1W^{-1}$:

$$\begin{aligned} af &= \frac{f_{11} \cdot H_{I_S^{nv}} + f_{13} \cdot a_S}{D_S^{nv}} \cdot \frac{f_{22} \cdot H_{I_S^v} + f_{24} \cdot a_S^v}{D_S^v} \\ &= (1 - V_{perc}) \frac{\beta_C \cdot H_{I_S^{nv}} + \beta_I \cdot a_S}{D_S^{nv}} \cdot V_{perc}(1 - v_a) \frac{\beta_C \cdot H_{I_S^v} + \beta_I \cdot a_S^v}{D_S^v} \\ &= V_{perc}(1 - v_a) \frac{\beta_C \cdot H_{I_S^{nv}} + \beta_I \cdot a_S}{D_S^{nv}} \cdot V_{perc}(1 - v_a) \frac{\beta_C \cdot H_{I_S^v} + \beta_I \cdot a_S^v}{D_S^v} \\ &= \frac{f_{21} \cdot H_{I_S^{nv}} + f_{23} \cdot a_S}{D_S^{nv}} \cdot \frac{f_{12} \cdot H_{I_S^v} + f_{14} \cdot a_S^v}{D_S^v} \\ &= eb \end{aligned}$$

Thus, the discriminant is $\Delta = (a+f)^2$. We conclude that the discriminant is positive, and we have in both cases two different eigenvalues and the max of them is: $\lambda = a + f$.

We denote $R_{0S}$ the reproductive number associated to sensitive infections which is the spectral radius of $F_1W^{-1}$ and $R_{0R}$ the one for resistant infections which equals the spectral radius of $F_2Y^{-1}$. We obtain:

$$\begin{aligned}
R_{0S} &= (1-V_{perc})\frac{\beta_C+\beta_I\frac{a_S}{\eta_S+\lambda_{A,I}+\gamma_{A,S}+\psi_A+\phi_{A,I}}}{\lambda_S+\lambda_{A,C}+\phi_{A,C}+a_S-\eta_S\frac{a_S}{\eta_S+\lambda_{A,I}+\gamma_{A,S}+\psi_A+\phi_{A,I}}} \\
&\quad + V_{perc}(1-v_a)\frac{\beta_C+\beta_I\frac{a_S(1-v_i)}{\frac{\eta_S}{1-v_r}+\lambda_{A,I}+\gamma_{A,S}+\psi_A+\phi_{A,I}}}{\frac{\lambda_S}{1-v_d}+\lambda_{A,C}+\phi_{A,C}+a_S(1-v_i)-\frac{\eta_S}{1-v_r}\frac{a_S(1-v_i)}{\frac{\eta_S}{1-v_r}+\lambda_{A,I}+\gamma_{A,S}+\psi_A+\phi_{A,I}}} \\
R_{0R} &= f(1-V_{perc})\frac{\beta_C+\beta_I\frac{a_R}{\eta_R+\gamma_{A,R}}}{\lambda_R+a_R-\eta_R\frac{a_R}{\eta_R+\gamma_{A,R}}}+fV_{perc}(1-v_a)\frac{\beta_C+\beta_I\frac{a_R(1-v_i)}{\frac{\eta_R}{1-v_r}+\gamma_{A,R}}}{\frac{\lambda_R}{1-v_d}+a_R(1-v_i)-\frac{\eta_R}{1-v_r}\frac{a_R(1-v_i)}{\frac{\eta_R}{1-v_r}+\gamma_{A,R}}} \\
R_0 &= \max(R_{0S}, R_{0R})
\end{aligned} \tag{A1}$$

In order to obtain $R_0$, we numerically compute the max of the two eigenvalues (the one from $F_1W^{-1}$ and the one from $F_2Y^{-1}$).

We can compute those eigenvalues for a particular case: the absence of a vaccine. It leads to $V_{perc} = 0$, thus we obtain:

$$\begin{aligned}
R_{0S} &= \frac{\beta_C+\beta_I\frac{a_S}{\eta_S+\lambda_{A,I}+\gamma_{A,S}+\psi_A+\phi_{A,I}}}{\lambda_S+\lambda_{A,C}+\phi_{A,C}+a_S-\eta_S\frac{a_S}{\eta_S+\lambda_{A,I}+\gamma_{A,S}+\psi_A+\phi_{A,I}}} \\
R_{0R} &= f\frac{\beta_C+\beta_I\frac{a_R}{\eta_R+\gamma_{A,R}}}{\lambda_R+a_R-\eta_R\frac{a_R}{\eta_R+\gamma_{A,R}}} \\
R_0 &= \max(R_{0S}, R_{0R})
\end{aligned} \tag{A2}$$

For a given vaccine type with associated vaccine efficacies, the minimum vaccine coverage needed to have $R_0$ below 1 depends on which one of $R_{0S}$ and $R_{0R}$ is higher.

If $R_{0S} > R_{0R}$, then the minimum vaccine coverage is given by

$$V^{\star}_{perc} = \frac{1-\frac{\beta_C+\beta_I\frac{a_S}{\eta_S+\lambda_{A,I}+\gamma_{A,S}+\psi_A+\phi_{A,I}}}{D_s^{nv}}}{(1-v_a)\frac{\beta_C+\beta_I\frac{a_S(1-v_i)}{\frac{\eta_S}{1-v_r}+\lambda_{A,I}+\gamma_{A,S}+\psi_A+\phi_{A,I}}}{D_s^{v}}-\frac{\beta_C+\beta_I\frac{a_S}{\eta_S+\lambda_{A,I}+\gamma_{A,S}+\psi_A+\phi_{A,I}}}{D_s^{nv}}}.$$

If $R_{0S} < R_{0R}$, then the minimum vaccine coverage is given by

$$V^{\star}_{perc} = \frac{1-f\frac{\beta_C+\beta_I\frac{a_R}{\eta_R+\gamma_{A,R}}}{\lambda_R+a_R-\eta_R\frac{a_R}{\eta_R+\gamma_{A,R}}}}{f(1-v_a)\frac{\beta_C+\beta_I\frac{a_R(1-v_i)}{\frac{\eta_R}{1-v_r}+\gamma_{A,R}}}{\frac{\lambda_R}{1-v_d}+a_R(1-v_i)-\frac{\eta_R}{1-v_r}\frac{a_R(1-v_i)}{\frac{\eta_R}{1-v_r}+\gamma_{A,R}}}-f\frac{\beta_C+\beta_I\frac{a_R}{\eta_R+\gamma_{A,R}}}{\lambda_R+a_R-\eta_R\frac{a_R}{\eta_R+\gamma_{A,R}}}}.$$

##### A.2 Model equilibria

For a given non-negative initial condition, the system has a unique non-negative maximal solution linearly dependant of the total population size. The unicity ensures that when no one is vaccinated (or, alternatively, everyone is vaccinated), the respective compartments stay null throughout time.

We analytically derived the equilibria of the model for various assumptions presented in Table 2. For all assumptions without vaccination, we remove the upper script $nv$ for better readability.

##### A.2.1 Assumption 1

For Assumption 1, all antibiotic-related parameters are set to zero. We search for the equilibria and solve the following system:

$$\begin{cases} 0 = \frac{dU}{dt} = -\frac{\beta_C C_S}{N}U - \frac{\beta_I I_S}{N}U - \frac{\beta_C f C_R}{N}U - \frac{\beta_I f I_R}{N}U + \frac{1}{d_S}C_S + \frac{1}{d_R}C_R \\ 0 = \frac{dC_S}{dt} = \frac{\beta_C C_S}{N}U + \frac{\beta_I I_S}{N}U - \frac{1}{d_S}C_S - a_S C_S + \eta_S I_S \\ 0 = \frac{dC_R}{dt} = \frac{\beta_C f C_R}{N}U + \frac{\beta_I f I_R}{N}U - \frac{1}{d_R}C_R - a_R C_R + \eta_R I_R \\ 0 = \frac{dC_S}{dt} = a_S C_S - \eta_S I_S \\ 0 = \frac{dI_R}{dt} = a_R C_R - \eta_R I_R \end{cases} \tag{A3}$$

The last two equations of (A3) are equivalent to $I_S = \frac{a_S}{\eta_S}C_S$ and $I_R = \frac{a_R}{\eta_R}C_R$.
The second and third equations of (A3) are equivalent to:

$$\begin{cases} 0 = C_S\left(\left(\beta_C + \beta_I\frac{a_S}{\eta_S}\right)\frac{U}{N} - \frac{1}{d_S}\right) \\ 0 = C_R\left(\left(\beta_C + \beta_I\frac{a_R}{\eta_R}\right)\frac{fU}{N} - \frac{1}{d_R}\right) \end{cases} \Leftrightarrow \begin{cases} C_S = 0 \text{ or } U = \frac{N}{d_S\left(\beta_C + \beta_I\frac{a_S}{\eta_S}\right)} \\ C_R = 0 \text{ or } U = \frac{N}{d_R f\left(\beta_C + \beta_I\frac{a_R}{\eta_R}\right)} \end{cases}.$$

Coexistence happens when $C_S \neq 0$ and $C_R \neq 0$. It is only possible if

$$U = \frac{N}{d_S\left(\beta_C + \beta_I\frac{a_S}{\eta_S}\right)} = \frac{N}{d_R f\left(\beta_C + \beta_I\frac{a_R}{\eta_R}\right)} \Leftrightarrow f = \frac{d_S\left(\beta_C + \beta_I\frac{a_S}{\eta_S}\right)}{d_R\left(\beta_C + \beta_I\frac{a_R}{\eta_R}\right)}.$$

Moreover, in order to have $U \leq N$ , we need $\left(\beta_C + \beta_I\frac{a_S}{\eta_S}\right)d_S \geq 1$.
All possible equilibria are:

- the disease-free equilibrium verifying $U = N$ and $C_S = C_R = I_S = I_R = 0$,
- the equilibrium with sensitive only verifying

$$U = \frac{N}{d_S\left(\beta_C + \beta_I\frac{a_S}{\eta_S}\right)}, C_R = I_R = 0, C_S = \frac{N\eta_S}{\eta_S + a_S}\left(1 - \frac{1}{d_S\left(\beta_C + \beta_I\frac{a_S}{\eta_S}\right)}\right), \text{ and}$$

$$I_S = \frac{N a_S}{\eta_S + a_S}\left(1 - \frac{1}{d_S\left(\beta_C + \beta_I\frac{a_S}{\eta_S}\right)}\right),$$

- the equilibrium with resistant only verifying

$$U = \frac{N}{d_R f\left(\beta_C + \beta_I\frac{a_R}{\eta_R}\right)}, C_S = I_S = 0, C_R = \frac{N\eta_R}{\eta_R + a_R}\left(1 - \frac{1}{d_R f\left(\beta_C + \beta_I\frac{a_R}{\eta_R}\right)}\right), \text{ and}$$

$$I_R = \frac{N a_R}{\eta_R + a_R}\left(1 - \frac{1}{d_R f\left(\beta_C + \beta_I\frac{a_R}{\eta_R}\right)}\right),$$

- the equilibrium with coexistence verifying

$$U = \frac{N}{d_S\left(\beta_C + \beta_I \frac{a_S}{\eta_S}\right)} = \frac{N}{d_R f\left(\beta_C + \beta_I \frac{a_R}{\eta_R}\right)}, N - U = C_S\left(1 + \frac{a_S}{\eta_S}\right) + C_R\left(1 + \frac{a_R}{\eta_R}\right).$$

##### A.2.2 Assumption 2

For Assumption 2, we only consider the fluxes of decolonisation due to antibiotics associated with bystander exposure or bacteria-specific exposure. We solve the following system:

$$\begin{cases} 0 = -\frac{\beta_C C_S + \beta_I I_S}{N}U - \frac{\beta_C C_R + \beta_I I_R}{N}fU + \frac{1}{d_S}C_S + \frac{1}{d_R}C_R \\ \quad +\lambda_{A,C}C_S + \gamma_{A,R}I_R + \lambda_{A,I}I_S + \gamma_{A,S}I_S \\ 0 = -\frac{\beta_C C_S + \beta_I I_S}{N}U - \frac{\beta_C C_R + \beta_I I_R}{N}fU - \frac{1}{d_S}C_S - \lambda_{A,C}C_S - a_S C_S + \eta_S I_S \\ 0 = \frac{\beta_C C_R + \beta_I I_R}{N}fU - \frac{1}{d_R}C_R - a_R C_R + \eta_R I_R \\ 0 = a_S C_S - \eta_S I_S - \lambda_{A,I}I_S - \gamma_{A,S}I_S \\ 0 = a_R C_R - \eta_R I_R - \gamma_{A,R}I_R \end{cases} \tag{A4}$$

We obtain from the last two equations $I_S = \frac{a_S}{\eta_S + \lambda_{A,I} + \gamma_{A,S}}C_S$ and $I_R = \frac{a_R}{\eta_R + \gamma_{A,R}}C_R$. The second and third equations of (A4) are equivalent to:

$$\begin{cases} 0 = C_S\left(\frac{U}{N}\left(\beta_C + \beta_I \frac{a_S}{\eta_S + \lambda_{A,I} + \gamma_{A,S}}\right) - \frac{1}{d_S} - \lambda_{A,C} - a_S + \eta_S \frac{a_S}{\eta_S + \lambda_{A,I} + \gamma_{A,S}}\right) \\ 0 = C_R\left(\frac{fU}{N}\left(\beta_C + \beta_I \frac{a_R}{\eta_R + \gamma_{A,R}}\right) - \frac{1}{d_R} - a_R + \eta_R \frac{a_R}{\eta_R + \gamma_{A,R}}\right) \end{cases}$$

$$\Leftrightarrow \begin{cases} C_S = 0 \text{ or } U = \frac{N}{\beta_C + \beta_I \frac{a_S}{\eta_S + \lambda_{A,I} + \gamma_{A,S}}}\left(\frac{1}{d_S} + \lambda_{A,C} + a_S - \eta_S \frac{a_S}{\eta_S + \lambda_{A,I} + \gamma_{A,S}}\right) \\ C_R = 0 \text{ or } U = \frac{N}{f\left(\beta_C + \beta_I \frac{a_R}{\eta_R + \gamma_{A,R}}\right)}\left(\frac{1}{d_R} + a_R - \eta_R \frac{a_R}{\eta_R + \gamma_{A,R}}\right) \end{cases}.$$

Coexistence can happen if and only if both equations for U are equal, which leads to:

$$f = \frac{\frac{1}{d_R} + a_R - \eta_R \frac{a_R}{\eta_R + \gamma_{A,R}}}{\frac{1}{d_S} + \lambda_{A,C} + a_S - \eta_S \frac{a_S}{\eta_S + \lambda_{A,I} + \gamma_{A,S}}} \frac{\beta_C + \beta_I \frac{a_S}{\eta_S + \lambda_{A,I} + \gamma_{A,S}}}{\beta_C + \beta_I \frac{a_R}{\eta_R + \gamma_{A,R}}}.$$

Like for Assumption 1, there exists a unique expression of $f$ allowing coexistence.

##### A.2.3 Assumption 3

In this case, we add the selection rates due to antibiotics. The equations are now:

$$\begin{cases} 0 = -\frac{\beta_C C_S + \beta_I I_S}{N}U - \frac{\beta_C C_R + \beta_I I_R}{N}fU + \frac{1}{d_S}C_S + \frac{1}{d_R}C_R \\ \quad + \lambda_{A,C}C_S + \gamma_{A,R}I_R + \lambda_{A,I}I_S + \gamma_{A,S}I_S \\ 0 = \frac{\beta_C C_S + \beta_I I_S}{N}U - \frac{1}{d_S}C_S - \lambda_{A,C}C_S - a_S C_S + \eta_S I_S - \phi_{A,C}C_S \\ 0 = \frac{\beta_C C_R + \beta_I I_R}{N}fU - \frac{1}{d_R}C_R - a_R C_R + \eta_R I_R + \phi_{A,C}C_S \\ 0 = a_S C_S - \eta_S I_S - \lambda_{A,I}I_S - \gamma_{A,S}I_S - \psi_A I_S - \phi_{A,I}I_S \\ 0 = a_R C_R - \eta_R I_R - \gamma_{A,R}I_R + \psi_A I_S + \phi_{A,I}I_S \end{cases} \tag{A5}$$

From the last two equations, we obtain

$$I_S = \frac{a_S}{\eta_S + \lambda_{A,I} + \gamma_{A,S} + \psi_A + \phi_{A,I}} C_S \text{ and } I_R = \frac{a_R}{\eta_R + \gamma_{A,R}} C_R + \frac{\psi_A + \phi_{A,I}}{\eta_R + \gamma_{A,R}} I_S.$$

The second equation of (A5) is equivalent to:

$$\begin{aligned} 0 &= C_S \left( \frac{U}{N} \left( \beta_C + \beta_I \frac{a_S}{H_{I_S}} \right) - \frac{1}{d_S} - \lambda_{A,C} - a_S + \eta_S \frac{a_S}{H_{I_S}} - \phi_{A,C} \right) \\ &\Leftrightarrow C_S = 0 \text{ or } U = \frac{N}{\beta_C + \beta_I \frac{a_S}{H_{I_S}}} \left( \frac{1}{d_S} + \lambda_{A,C} + a_S - \eta_S \frac{a_S}{H_{I_S}} + \phi_{A,C} \right) \end{aligned}$$

where $H_{I_S} = \eta_S + \lambda_{A,I} + \gamma_{A,S} + \psi_A + \phi_{A,I}$.

To have $C_S \neq 0$, we need $U = \frac{N}{\beta_C + \beta_I \frac{a_S}{H_{I_S}}} \left( \frac{1}{d_S} + \lambda_{A,C} + a_S - \eta_S \frac{a_S}{H_{I_S}} + \phi_{A,C} \right)$. This is a valid expression if $U \geq 0$ and $U < N$. $U \geq 0$ is true, so the condition is:

$$U < N \Leftrightarrow \beta_C + \beta_I \frac{a_S}{H_{I_S}} > \frac{1}{d_S} + \lambda_{A,C} + a_S - \eta_S \frac{a_S}{H_{I_S}} + \phi_{A,C} \tag{A6}$$

Then the third equation of (A5) gives:

$$\begin{aligned} 0 = {} & C_R \left( \frac{fU}{N} \left( \beta_C + \beta_I \frac{a_R}{\eta_R + \gamma_{A,R}} \right) - \frac{1}{d_R} - a_R + \eta_R \frac{a_R}{\eta_R + \gamma_{A,R}} \right) \\ & + C_S \left( \left( \frac{\beta_I f U}{N} + \eta_R \right) \frac{\psi_A + \phi_{A,I}}{\eta_R + \gamma_{A,R}} \frac{a_S}{H_{I_S}} + \phi_{A,C} \right) \\ \Leftrightarrow C_S = {} & - \frac{\frac{fU}{N} \left( \beta_C + \beta_I \frac{a_R}{\eta_R + \gamma_{A,R}} \right) - \frac{1}{d_R} - a_R + \eta_R \frac{a_R}{\eta_R + \gamma_{A,R}}}{\left( \frac{\beta_I f U}{N} + \eta_R \right) \frac{\psi_A + \phi_{A,I}}{\eta_R + \gamma_{A,R}} \frac{a_S}{H_{I_S}} + \phi_{A,C}} C_R \\ \Leftrightarrow C_S = {} & - \frac{f \frac{\beta_C + \beta_I \frac{a_R}{\eta_R + \gamma_{A,R}}}{\beta_C + \beta_I \frac{a_s}{H_{I_S}}} \left( \frac{1}{d_S} + \lambda_{A,C} + a_S - \eta_S \frac{a_S}{H_{I_S}} + \phi_{A,C} \right) - \frac{1}{d_R} - a_R + \eta_R \frac{a_R}{\eta_R + \gamma_{A,R}}}{\left( \frac{\beta_I f}{\beta_C + \beta_I \frac{a_S}{H_{I_S}}} \left( \frac{1}{d_S} + \lambda_{A,C} + a_S - \eta_S \frac{a_S}{H_{I_S}} + \phi_{A,C} \right) + \eta_R \right) \frac{\psi_A + \phi_{A,I}}{\eta_R + \gamma_{A,R}} \frac{a_S}{H_{I_S}} + \phi_{A,C}} C_R \\ \Leftrightarrow C_S = {} & -E C_R. \end{aligned}$$

In order to have $C_R > 0$ and $C_S > 0$, we need

$$E < 0 \Leftrightarrow f < \frac{\frac{1}{d_R} + a_R - \eta_R \frac{a_R}{\eta_R + \gamma_{A,R}}}{\frac{1}{d_S} + \lambda_{A,C} + a_S - \eta_S \frac{a_S}{H_{I_S}} + \phi_{A,C}} \frac{\beta_C + \beta_I \frac{a_S}{H_{I_S}}}{\beta_C + \beta_I \frac{a_R}{\eta_R + \gamma_{A,R}}}. \tag{A7}$$

Now we try to obtain the exact value of each compartment. For this we use the fact that the population is constant:

$$\begin{aligned} & N = U + C_S + C_R + I_S + I_R \\ & \Leftrightarrow N = U - E C_R + C_R + I_S + \frac{a_R}{\eta_R + \gamma_{A,R}} C_R + \frac{\psi_A + \phi_{A,I}}{\eta_R + \gamma_{A,R}} I_S \\ & \Leftrightarrow N = U - E C_R + C_R - \frac{a_S}{H_{I_S}} E C_R \left( 1 + \frac{\psi_A + \phi_{A,I}}{\eta_R + \gamma_{A,R}} \right) + \frac{a_R}{\eta_R + \gamma_{A,R}} C_R \\ & \Leftrightarrow C_R \left( 1 + \frac{a_R}{\eta_R + \gamma_{A,R}} - E \left( 1 + \frac{a_S}{H_{I_S}} \left( 1 + \frac{\psi_A + \phi_{A,I}}{\eta_R + \gamma_{A,R}} \right) \right) \right) = N - U \\ & \Leftrightarrow C_R = \frac{N - U}{G} \end{aligned}$$

with

$$G = 1 + \frac{a_R}{\eta_R + \gamma_{A,R}} - E\left(1 + \frac{a_S}{\eta_S + \lambda_{A,I} + \gamma_{A,S} + \psi_A + \phi_{A,I}}\left(1 + \frac{\psi_A + \phi_{A,I}}{\eta_R + \gamma_{A,R}}\right)\right).$$

Due to the fact that we need $E < 0$, we have $G > 0$ and $C_R > 0$. To have $C_R < N$, we need

$$\begin{aligned}
&\frac{1}{G}\left[1 - \frac{1}{\beta_C + \beta_I \frac{a_S}{H_{I_S}}}\left(\frac{1}{d_S} + \lambda_{A,C} + a_S - \eta_S \frac{a_S}{H_{I_S}} + \phi_{A,C}\right)\right] < 1 \\
&\Leftrightarrow 1 - \frac{1}{\beta_C + \beta_I \frac{a_S}{H_{I_S}}}\left(\frac{1}{d_S} + \lambda_{A,C} + a_S - \eta_S \frac{a_S}{H_{I_S}} + \phi_{A,C}\right) \\
&\qquad < 1 + \frac{a_R}{\eta_R + \gamma_{A,R}} - E\left(1 + \frac{a_S}{H_{I_S}}\left(1 + \frac{\psi_A + \phi_{A,I}}{\eta_R + \gamma_{A,R}}\right)\right) \\
&\Leftrightarrow 0 < \frac{1}{\beta_C + \beta_I \frac{a_S}{H_{I_S}}}\left(\frac{1}{d_S} + \lambda_{A,C} + a_S - \eta_S \frac{a_S}{H_{I_S}} + \phi_{A,C}\right) \\
&\qquad + \frac{a_R}{\eta_R + \gamma_{A,R}} - E\left(1 + \frac{a_S}{H_{I_S}}\left(1 + \frac{\psi_A + \phi_{A,I}}{\eta_R + \gamma_{A,R}}\right)\right).
\end{aligned}$$

This last inequality is true, so $C_R < N$ without additional condition. To ensure $C_S < N$ we need

$$\begin{aligned}
&-E\frac{1}{G}\left[1 - \frac{1}{\beta_C + \beta_I \frac{a_S}{H_{I_S}}}\left(\frac{1}{d_S} + \lambda_{A,C} + a_S - \eta_S \frac{a_S}{H_{I_S}} + \phi_{A,C}\right)\right] < 1 \\
&\Leftrightarrow 1 - \frac{1}{\beta_C + \beta_I \frac{a_S}{H_{I_S}}}\left(\frac{1}{d_S} + \lambda_{A,C} + a_S - \eta_S \frac{a_S}{H_{I_S}} + \phi_{A,C}\right) \\
&\qquad < \frac{1}{-E}\left(1 + \frac{a_R}{\eta_R + \gamma_{A,R}} - E\left(1 + \frac{a_S}{H_{I_S}}\left(1 + \frac{\psi_A + \phi_{A,I}}{\eta_R + \gamma_{A,R}}\right)\right)\right) \\
&\Leftrightarrow {\color{red}\cancel{1}} - \frac{1}{\beta_C + \beta_I \frac{a_S}{H_{I_S}}}\left(\frac{1}{d_S} + \lambda_{A,C} + a_S - \eta_S \frac{a_S}{H_{I_S}} + \phi_{A,C}\right) \\
&\qquad < \frac{1}{-E}\left(1 + \frac{a_R}{\eta_R + \gamma_{A,R}}\right) + {\color{red}\cancel{1}} + \frac{a_S}{H_{I_S}}\left(1 + \frac{\psi_A + \phi_{A,I}}{\eta_R + \gamma_{A,R}}\right) \\
&0 < \frac{1}{\beta_C + \beta_I \frac{a_S}{H_{I_S}}}\left(\frac{1}{d_S} + \lambda_{A,C} + a_S - \eta_S \frac{a_S}{H_{I_S}} + \phi_{A,C}\right) \\
&\qquad + \frac{1}{-E}\left(1 + \frac{a_R}{\eta_R + \gamma_{A,R}}\right) + \frac{a_S}{H_{I_S}}\left(1 + \frac{\psi_A + \phi_{A,I}}{\eta_R + \gamma_{A,R}}\right)
\end{aligned}$$

This last inequality is true, so $C_S < N$.

Finally, we verify:

$$\begin{aligned}
&I_S < N \\
&\Leftrightarrow 1 - \frac{1}{\beta_C + \beta_I \frac{a_S}{H_{I_S}}}\left(\frac{1}{d_S} + \lambda_{A,C} + a_S - \eta_S \frac{a_S}{H_{I_S}} + \phi_{A,C}\right) \\
&\qquad < {\color{red}\frac{1 + \frac{a_S}{H_{I_S}}\left(1 + \frac{\psi_A + \phi_{A,I}}{\eta_R + \gamma_{A,R}}\right)}{\frac{a_S}{H_{I_S}}}} - \frac{1}{E\frac{a_S}{H_{I_S}}}\left(1 + \frac{a_R}{\eta_R + \gamma_{A,R}}\right)
\end{aligned}$$

Yet the term in red is greater than 1 thus the inequality is true.

We have proved that under Conditions (A6) and (A7), all compartments have their value between 0 and $N$ which corresponds to acceptable values.

If Condition (A7) does not hold and $E = 0$, only resistant strains survive with $C_R = \frac{N-U}{1+\frac{a_R}{\eta_R+\gamma_{A,R}}}$. $E$ being positive forces $C_S = C_R = 0$ because they are population numbers. If Condition (A6) does not hold and becomes an equality, then uncolonised individuals form the whole population and bacterial strains do not exist. If the inequality is reversed, it forces $C_S$ to zero which impedes coexistence of both strains. This concludes the study of Assumption 3.

##### A.2.4 Assumption 4

We consider that the entire population is vaccinated and we compute the equilibria to compare them with Assumption 1. The system to solve is the following:

$$\begin{cases} 0 = \frac{dU^v}{dt} = -\frac{\beta_C C_S^v + \beta_I I_S^v}{N}(1-v_a)U^v - \frac{\beta_C C_R^v + \beta_I I_R^v}{N} f(1-v_a)U^v \\ \qquad + \frac{1}{d_S(1-v_d)}C_S^v + \frac{1}{d_R(1-v_d)}C_R^v \\ 0 = \frac{dC_S^v}{dt} = \frac{\beta_C C_S^v + \beta_I I_S^v}{N}(1-v_a)U^v - \frac{1}{d_S(1-v_d)}C_S^v - a_S(1-v_i)C_S^v + \frac{\eta_S}{(1-v_r)}I_S^v \\ 0 = \frac{dC_R^v}{dt} = \frac{\beta_C C_R^v + \beta_I I_R^v}{N} f(1-v_a)U^v - \frac{1}{d_R(1-v_d)}C_R^v - a_R(1-v_i)C_R^v + \frac{\eta_R}{(1-v_r)}I_R^v \\ 0 = \frac{dI_S^v}{dt} = a_S(1-v_i)C_S^v - \frac{\eta_S}{(1-v_r)}I_S^v \\ 0 = \frac{dI_R^v}{dt} = a_R(1-v_i)C_R^v - \frac{\eta_R}{(1-v_r)}I_R^v \end{cases} \tag{A8}$$

From the last two equations we obtain $I_S^v = \frac{a_S(1-v_i)(1-v_r)}{\eta_S}C_S^v$ and $I_R^v = \frac{a_R(1-v_i)(1-v_r)}{\eta_R}C_R^v$. The second and third equations of the system (A8) are equivalent to:

$$\begin{cases} 0 = C_S^v \left( \frac{(1-v_a)U^v}{N} \left( \beta_C + \beta_I \frac{a_S(1-v_i)(1-v_r)}{\eta_S} \right) - \frac{1}{d_S(1-v_d)} \right) \\ 0 = C_R^v \left( \frac{f(1-v_a)U^v}{N} \left( \beta_C + \beta_I \frac{a_R(1-v_i)(1-v_r)}{\eta_R} \right) - \frac{1}{d_R(1-v_d)} \right) \end{cases}$$

$$\Leftrightarrow \begin{cases} C_S^v = 0 \text{ or } U^v = \frac{N}{\left(\beta_C + \beta_I \frac{a_S(1-v_i)(1-v_r)}{\eta_S}\right)(1-v_a)d_S(1-v_d)} \\ C_R^v = 0 \text{ or } U^v = \frac{N}{\left(\beta_C + \beta_I \frac{a_R(1-v_i)(1-v_r)}{\eta_R}\right) f(1-v_a)d_R(1-v_d)} \end{cases}.$$

In order to have coexistence, we need both expressions of $U^v$ to be equal, which is equivalent to:

$$f = \frac{d_S}{d_R} \frac{\beta_C + \beta_I \frac{a_S(1-v_i)(1-v_r)}{\eta_S}}{\beta_C + \beta_I \frac{a_R(1-v_i)(1-v_r)}{\eta_R}}.$$

In this case we have a unique value of $f$ that allows coexistence. The condition is modified by $v_i$ and $v_r$.

We also need $U^v \leq N$ and it is true if and only if

$$\left(\beta_C + \beta_I \frac{a_S(1-v_i)(1-v_r)}{\eta_S}\right)(1-v_a)d_S(1-v_d) \geq 1.$$

The condition for which the coexistence equilibrium exists is also different than before: in fact, if vaccine parameters are sufficiently reducing the left part of the inequality then a previous thriving bacteria could be eliminated by the vaccine at equilibrium.

##### A.2.5 Assumption 5

We consider that the entire population is vaccinated and we compute the equilibria to compare them with Assumption 2. We solve the following system:

$$\begin{cases} 0 = -\dfrac{\beta_C C_S^v + \beta_I I_S^v}{N}(1-v_a)U^v - \dfrac{\beta_C C_R^v + \beta_I I_R^v}{N} f(1-v_a)U^v \\ \quad + \dfrac{1}{d_S(1-v_d)}C_S^v + \dfrac{1}{d_R(1-v_d)}C_R^v + \lambda_{A,C}C_S^v + \gamma_{A,R}I_R^v + \lambda_{A,I}I_S^v + \gamma_{A,S}I_S^v \\ 0 = \dfrac{\beta_C C_S^v + \beta_I I_S^v}{N}(1-v_a)U^v - \dfrac{1}{d_S(1-v_d)}C_S^v - a_S(1-v_i)C_S^v + \dfrac{\eta_S}{(1-v_r)}I_S^v - \lambda_{A,C}C_S^v \\ 0 = \dfrac{\beta_C C_R^v + \beta_I I_R^v}{N} f(1-v_a)U^v - \dfrac{1}{d_R(1-v_d)}C_R^v - a_R(1-v_i)C_R^v + \dfrac{\eta_R}{(1-v_r)}I_R^v \\ 0 = a_S(1-v_i)C_S^v - \dfrac{\eta_S}{(1-v_r)}I_S^v - \lambda_{A,I}I_S^v - \gamma_{A,S}I_S \\ 0 = a_R(1-v_i)C_R^v - \dfrac{\eta_R}{(1-v_r)}I_R^v - \gamma_{A,R}I_R^v \end{cases} \tag{A9}$$

By analogy with Assumption 2, we obtain a unique value of $f$ allowing coexistence between sensitive and resistant strains:

$$f = \frac{\frac{1}{d_R(1-v_d)} + a_R(1-v_i) - \frac{\eta_R}{(1-v_r)}\frac{a_R(1-v_i)}{\frac{\eta_R}{(1-v_r)}+\gamma_{A,R}}}{\frac{1}{d_S(1-v_d)} + \lambda_{A,C} + a_S(1-v_i) - \frac{\eta_S}{(1-v_r)}\frac{a_S(1-v_i)}{\frac{\eta_S}{(1-v_r)}+\lambda_{A,I}+\gamma_{A,S}}} \frac{\beta_C + \beta_I\frac{a_S(1-v_i)}{\frac{\eta_S}{1-v_r}+\lambda_{A,I}+\gamma_{A,S}}}{\beta_C + \beta_I\frac{a_R(1-v_i)}{\frac{\eta_R}{1-v_r}+\gamma_{A,R}}}$$

This means that if both strains can coexist thanks to a certain relative fitness, the introduction of a vaccine can disrupt the coexistence. Two things can happen: either one or the other strains dominate, or, if the vaccine is strong enough, both strains disappear. Reciprocally, if both strains can't coexist and a vaccine is introduced before total disappearance of the dominated strain, coexistence is created if the relative fitness now equals the formula given just above.

##### A.2.6 Assumption 6

We consider that the entire population is vaccinated and both decolonisation and selection fluxes due to antibiotics are used. The system to solve is:

$$\begin{cases} 0 = -\dfrac{\beta_C C_S^v + \beta_I I_S^v}{N}(1-v_a)U^v - \dfrac{\beta_C C_R^v + \beta_I I_R^v}{N} f(1-v_a)U^v \\ \quad + \dfrac{1}{d_S(1-v_d)}C_S^v + \dfrac{1}{d_R(1-v_d)}C_R^v + \lambda_{A,C}C_S^v + \gamma_{A,R}I_R^v + \lambda_{A,I}I_S^v + \gamma_{A,S}I_S^v \\ 0 = \dfrac{\beta_C C_S^v + \beta_I I_S^v}{N}(1-v_a)U^v - \dfrac{1}{d_S(1-v_d)}C_S^v - a_S(1-v_i)C_S^v + \dfrac{\eta_S}{(1-v_r)}I_S^v \\ \quad - \lambda_{A,C}C_S^v - \phi_{A,C}C_S^v \\ 0 = \dfrac{\beta_C C_R^v + \beta_I I_R^v}{N} f(1-v_a)U^v - \dfrac{1}{d_R(1-v_d)}C_R^v - a_R(1-v_i)C_R^v + \dfrac{\eta_R}{(1-v_r)}I_R^v + \phi_{A,C}C_S^v \\ 0 = a_S(1-v_i)C_S^v - \dfrac{\eta_S}{(1-v_r)}I_S^v - \lambda_{A,I}I_S^v - \gamma_{A,S}I_S - \phi_{A,I}I_S^v - \psi_A I_S^v \\ 0 = a_R(1-v_i)C_R^v - \dfrac{\eta_R}{(1-v_r)}I_R^v - \gamma_{A,R}I_R^v + \phi_{A,I}I_S^v + \psi_A I_S^v \end{cases} \tag{A10}$$

By analogy with Assumption 3, coexistence between both strains is possible under Conditions A11 and A12.

$$(1-v_a)\left(\beta_C+\beta_I\frac{a_S(1-v_i)}{\frac{\eta_S}{1-v_r}+\lambda_{A,I}+\gamma_{A,S}+\psi_A+\phi_{A,I}}\right)$$
$$>\frac{1}{d_S(1-v_d)}+\lambda_{A,C}+a_S(1-v_i)-\frac{\eta_S}{(1-v_r)}\frac{a_S(1-v_i)}{\frac{\eta_S}{(1-v_r)}+\lambda_{A,I}+\gamma_{A,S}+\psi_A+\phi_{A,I}}+\phi_{A,C} \tag{A11}$$

$$f<\frac{\frac{1}{d_R(1-v_d)}+\frac{a_R(1-v_i)\gamma_{A,R}}{\frac{\eta_R}{(1-v_r)}+\gamma_{A,R}}}{\frac{1}{d_S(1-v_d)}+\lambda_{A,C}+\frac{a_S(1-v_i)(\lambda_{A,I}+\gamma_{A,S}+\psi_A+\phi_{A,I})}{\frac{\eta_S}{(1-v_r)}+\lambda_{A,I}+\gamma_{A,S}+\psi_A+\phi_{A,I}}+\phi_{A,C}}\frac{\beta_C+\beta_I\frac{a_S(1-v_i)}{\frac{\eta_S}{(1-v_r)}+\lambda_{A,I}+\gamma_{A,S}+\psi_A+\phi_{A,I}}}{\beta_C+\beta_I\frac{a_R(1-v_i)}{\frac{\eta_R}{1-v_r}+\gamma_{A,R}}} \tag{A12}$$

If both strains coexist and a vaccine is introduced, there are several possibilities. If both equations (A11) and (A12) are true, then there is still coexistence with the following expressions of the compartments.

$$U^v=\frac{N}{\beta_C^v+\beta_I^v\frac{a_S^v}{\eta_s^v+\lambda_{A,I}+\gamma_{A,S}+\psi_A+\phi_{A,I}}}\left(\frac{1}{d_S^v}+\lambda_{A,C}+a_S^v-\eta_S^v\frac{a_S^v}{\eta_S^v+\lambda_{A,I}+\gamma_{A,S}+\psi_A+\phi_{A,I}}+\phi_{A,C}\right)$$

$$C_S^v=-\frac{\frac{fU^v}{N}\left(\beta_C^v+\beta_I^v\frac{a_R^v}{\eta_R^v+\gamma_{A,R}}\right)-\frac{1}{d_R^v}-a_R^v+\eta_R^v\frac{a_R^v}{\eta_R^v+\gamma_{A,R}}}{\left(\frac{\beta_I^v fU}{N}+\eta_R^v\right)\frac{\psi_A+\phi_{A,I}}{\eta_R^v+\gamma_{A,R}}\frac{a_S^v}{\eta_S^v+\lambda_{A,I}+\gamma_{A,S}+\psi_A+\phi_{A,I}}+\phi_{A,C}}C_R^v=-E^vC_R^v$$

$$C_R^v=\frac{N-U^v}{1+\frac{a_R^v}{\eta_R^v+\gamma_{A,R}}-E^v\left(1+\frac{a_S^v}{\eta_S^v+\lambda_{A,I}+\gamma_{A,S}+\psi_A+\phi_{A,I}}\left(1+\frac{\psi_A+\phi_{A,I}}{\eta_R^v+\gamma_{A,R}}\right)\right)}$$

with the notations: $\beta_C^v=\beta_C(1-v_a)$,$\beta_I^v=\beta_I(1-v_a)$, $d_X^v=d_X(1-v_d)$, $a_X^v=a_X(1-v_i)$, $\eta_X^v=\frac{\eta_X}{1-v_r}$.

If (A11) isn't true, then it forces $C_S^v=0=I_S^v$ and the third equation of (A10) gives

$$0=C_R^v\left(\frac{fU^v}{N}\left(\beta_C^v+\beta_I^v\frac{a_R^v}{\eta_R^v+\gamma_{A,R}}\right)-\frac{1}{d_R^v}-a_R^v+\eta_R^v\frac{a_R^v}{\eta_R^v+\gamma_{A,R}}\right)$$

which is equivalent to

$$C_R^v=0 \text{ or } U^v=\frac{N}{\left(\beta_C^v+\beta_I^v\frac{a_R^v}{\eta_R^v+\gamma_{A,R}}\right)f}\left(\frac{1}{d_R^v}+a_R^v-\eta_R^v\frac{a_R^v}{\eta_R^v+\gamma_{A,R}}\right).$$

And we have $C_R^v\neq 0$ if and only if

$$\left(\beta_C^v+\beta_I^v\frac{a_R^v}{\eta_R^v+\gamma_{A,R}}\right)f>\frac{1}{d_R^v}+a_R^v-\eta_R^v\frac{a_R^v}{\eta_R^v+\gamma_{A,R}}. \tag{A13}$$

More precisely, we have:

$$\begin{aligned}
&N=U^v+C_R^v+I_R^v\\
&\Leftrightarrow N=\frac{N}{\left(\beta_C^v+\beta_I^v\frac{a_R^v}{\eta_R^v+\gamma_{A,R}}\right)f}\left(\frac{1}{d_R^v}+a_R^v-\eta_R^v\frac{a_R^v}{\eta_R^v+\gamma_{A,R}}\right)+C_R^v\left(1+\frac{a_R^v}{\eta_R^v+\gamma_{A,R}}\right)\\
&\Leftrightarrow C_R^v=\frac{N-U^v}{1+\frac{a_R^v}{\eta_R^v+\gamma_{A,R}}}
\end{aligned}$$

$C_R^v$ is positive and less than $N$ if (A13) is true, in that case, the vaccine induces a resistant dominant strain.

If (A12) isn't true and (A11) still stands, then $E^v = 0$ which still forces $C_S^v = 0 = I_S^v$ and the vaccine still induces a resistant dominant strain.

The last possibility is the extinction of both strains when both conditions (A11) and (A13) aren't met.

#### A.3 Parameter value tables

##### A.3.1 Sensitivity analysis parameter tables

| Symbol | Description | Unit | Min | Max | Source |
|---|---|---|---|---|---|
| $\beta_C$ | Transmission rate from colonised | $\text{day}^{-1}$ | 0.01 | 0.1 | [3, 4, 5, 6] |
| $f$ | Relative fitness for transmission | | 0.9 | 1 | [4, 3, 7] |
| $a$ | Transition rate from colonised to infected | $\text{day}^{-1}$ | $5 \times 10^{-6}$ | $1 \times 10^{-4}$ | [4, 5, 7] |
| $\tau_{rec}$ | Time until infection recovery without antibiotic | day | 10 | 50 | assumed |
| $d$ | Colonisation duration | day | 30 | 130 | [3, 8, 9] |
| $p_{by}$ | Proportion of bystander exposure | | 0.001 | 0.01 | [10] |
| $\tau_{dec,by}$ | Time until clearance by bystander antibiotic | day | 10 | 30 | assumed |
| $p_{min,C}$ | Proportion of colonised individuals by sensitive strain with a minority resistant strain | | 0 | 0.5 | assumed |
| $p_{sp,s}$ | Proportion of infected individuals by sensitive strain taking specific antibiotics | | 0.5 | 1 | assumed |
| $\tau_{dec,sp,s}$ | Time until clearance by specific antibiotic for the sensitive strain | day | 3 | 15 | assumed |
| $p_{min,I}$ | Proportion of infected individuals by sensitive strain with a minority resistant strain | | 0 | 0.5 | assumed |
| $p_{sp,r}$ | Proportion of infected individuals by resistant strain taking specific antibiotics | | 0.5 | 1 | assumed |
| $\tau_{dec,sp,r}$ | Time until clearance by specific antibiotic for the resistant strain | day | 5 | 40 | assumed |

Table A1: **Parameter ranges used for sensitivity analysis** (for each parameter, a uniform distribution was considered, all parameter not mentioned were fixed to 0). All vaccine parameters were ranged from 0.3 to 0.9, vaccine coverage was ranged from 10% to 90%.

##### A.3.2 Parameters of two example bacteria

| Symbol | Description | Value | Source |
|---|---|---|---|
| $\beta_C$ | Transmission rate from colonised ($\text{days}^{-1}$) | $1.53e^{-2}$ | estimated |
| $\beta_I$ | Transmission rate from infected ($\text{days}^{-1}$) | 0 | assumed |
| $f$ | Relative fitness for transmission | 0.92 | estimated |
| $d$ | Colonisation duration (days) | 98 | [7] |
| $a$ | Transition rate from colonised to infected for sensitive strain ($\text{days}^{-1}$) | $3.46e^{-6}$ | estimated |
| $\tau_{rec}$ | Time until infection recovery without antibiotic (days) | 50 | assumed |
| $p_{by}$ | Proportion of bystander exposure | $4.76e^{-3}$ | [11] |
| $\tau_{dec,by}$ | Time until clearance by bystander antibiotic (days) | 10 | assumed |
| $p_{min,C}$ | Proportion of colonised individuals by sensitive strain with a minority resistant strain | 0.1 | [12] |
| $p_{sp,s}$ | Proportion of infected individuals by sensitive strain taking specific antibiotics | 1 | assumed |
| $\tau_{dec,sp,s}$ | Time until clearance by specific antibiotic for the sensitive strain (days) | 14 | [13] |
| $p_{min,I}$ | Proportion of infected individuals by sensitive strain with a minority resistant strain | 0 | assumed |
| $p_{sp,r}$ | Proportion of infected individuals by resistant strain taking specific antibiotics | 1 | assumed |
| $\tau_{dec,sp,r}$ | Time until clearance by specific antibiotic for the resistant strain (days) | 14 | [13] |

Table A2: Parameter symbols, descriptions and values for *Staphylococcus aureus*.

| Symbol | Description | Value | Source |
|---|---|---|---|
| $\beta_C$ | Transmission rate from colonised (days$^{-1}$) | 0.062 | estimated |
| $\beta_I$ | Transmission rate from infected (days$^{-1}$) | 0.062 | estimated |
| $f$ | Relative fitness for transmission | 0.855 | estimated |
| $d$ | Colonisation duration (days) | 365 | assumed |
| $a$ | Transition rate from colonised to infected (days$^{-1}$) | $1.684e^{-4}$ | estimated |
| $\tau_{rec}$ | Time until infection recovery without antibiotic (days) | 28 | assumed |
| $p_{by}$ | Proportion of bystander exposure | $1.73e^{-3}$ | [11] |
| $\tau_{dec,by}$ | Time until clearance by bystander antibiotic (days) | 10 | assumed |
| $p_{min,C}$ | Proportion of colonised individuals by sensitive strain with a minority resistant strain | 0.1 | assumed |
| $p_{sp,s}$ | Proportion of infected individuals by sensitive strain taking specific antibiotics | 0.8 | assumed |
| $p_{sp,r}$ | Proportion of infected individuals by resistant strain taking specific antibiotics | 0.8 | assumed |
| $\gamma_{A,S}$ | Rate of clearance by specific antibiotic against sensitive strains (days$^{-1}$) | 0.326 | See 2.4.2 |
| $\psi_A$ | Rate of selection by specific antibiotic against sensitive strains (days$^{-1}$) | $2.286e^{-3}$ | See 2.4.2 |
| $\gamma_{A,R}$ | Rate of clearance by specific antibiotic against resistant strains (days$^{-1}$) | 0.322 | See 2.4.2 |

Table A3: Parameter symbols, descriptions and values for *Escherichia coli*.

#### A.4 Supplementary figures

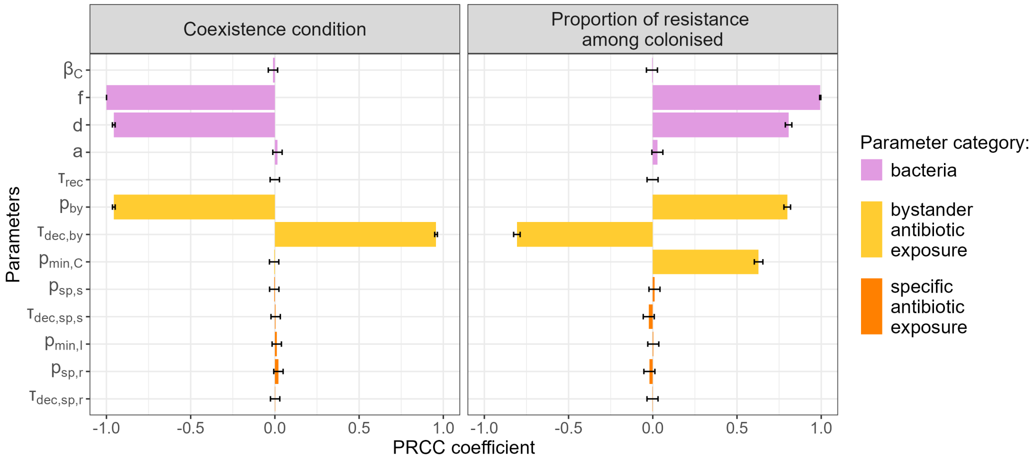

(a) For *S. aureus*

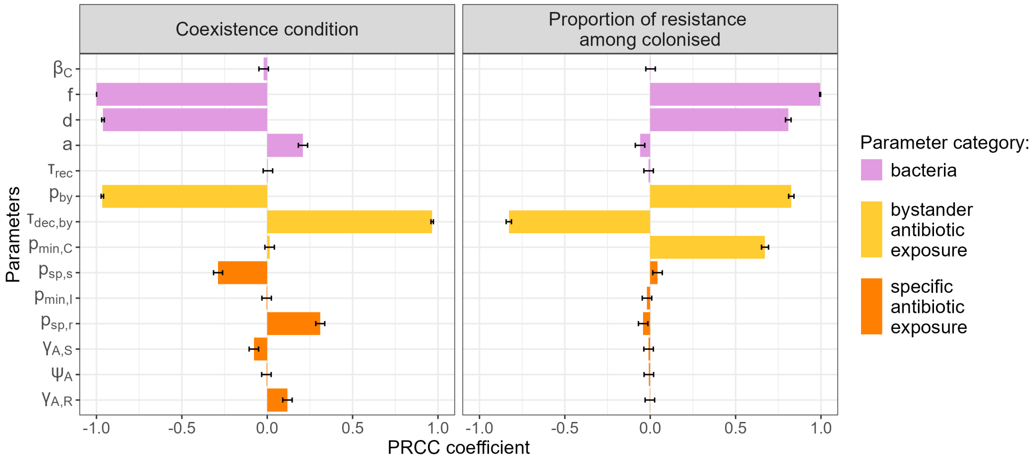

(b) For *E. coli*

Figure A1: **Partial rank correlation coefficients (PRCC) for the coexistence condition and the proportion of resistant colonisation among colonised at equilibrium.** Parameter ranges are at more or less 10% of the values in tables A2 and A3. The coexistence condition was transformed to a continuous variable by taking the right-hand side minus the left-hand side of the equation (A7). Coexistence is satisfied if this new variable is positive. The lower this variable is, the greater the risk that it will be negative, which corresponds to the coexistence condition not being met. Here the relative fitness $f$ is the most influential parameter. For the set of parameters with coexistence, we observed the impact of parameters on the proportion of resistant colonised, here the relative fitness remains the most influential parameter.

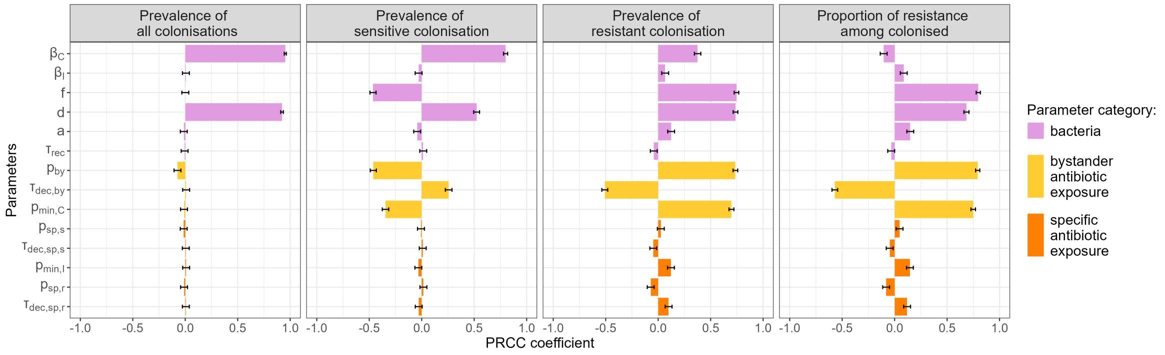

Figure A2: **Partial rank correlation coefficients (PRCC) for different outputs of a simulation until equilibrium without vaccine.** Parameter ranges considered are presented in Table A1 and explored through Latin Hypercube Sampling (5 000 samples). PRCC indicates the monotony between a specific parameter and an output variable. Positive sign corresponds to an increase, negative sign to a decrease.

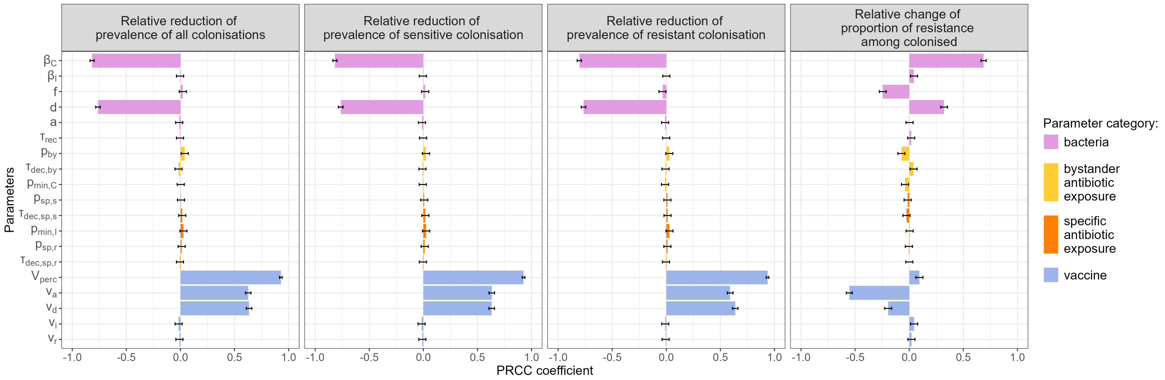

Figure A3: **Partial rank correlation coefficients (PRCC) for different outputs of the comparison between one year simulations without vaccine and with vaccine.** Parameter ranges considered are presented in Table A1 and explored through Latin Hypercube Sampling (5 000 samples). PRCC indicates the monotony between a specific parameter and an output variable. Positive sign corresponds to an increase, negative sign to a decrease.

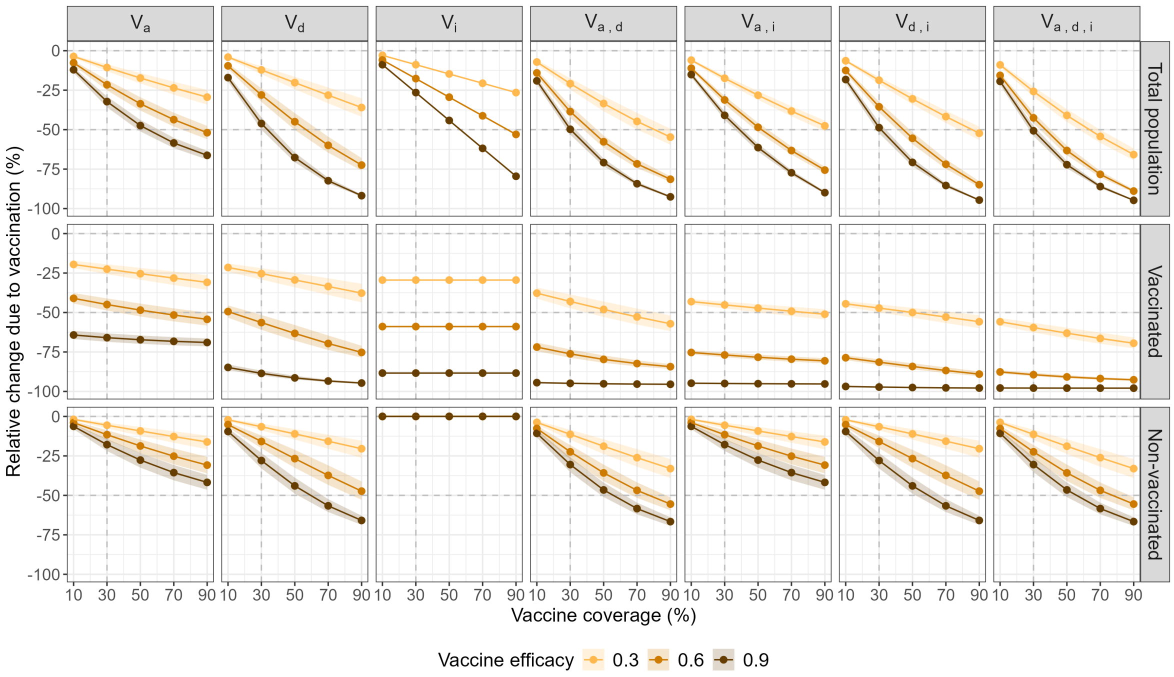

(a) For *S.aureus*

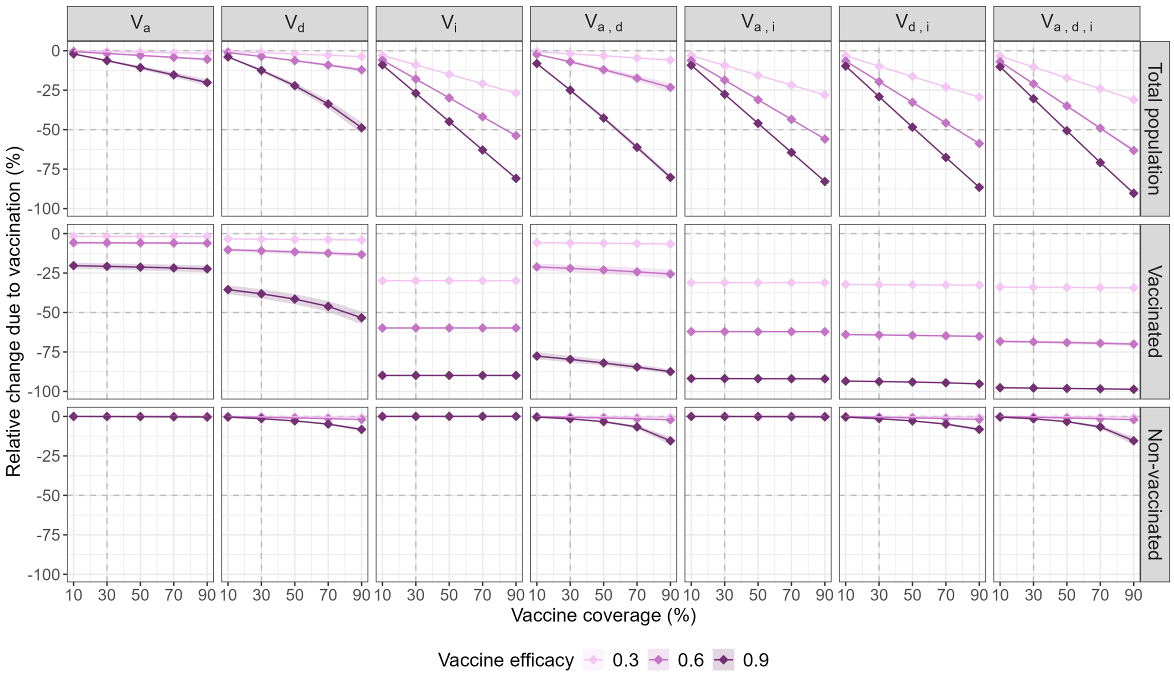

(b) For *E.coli*

Figure A4: **Impact of vaccines on the annual cumulative incidence of resistant infections for two bacteria.** The points indicate the median relative reduction in the predicted incidence of resistant infections as a function of vaccine coverage, as compared with the no-vaccine incidence, and the ribbon depicts the 95% interval between the 2.5th and 97.5th percentiles. Each column corresponds to one vaccine type defined in Table 1. For each considered vaccine, three levels of efficacy are considered: 30% (light), 60% (medium) and 90% (dark), assuming identical vaccine-driven impacts on model parameters for vaccines that incorporate several. Each row corresponds to a specific population (total, vaccinated or non-vaccinated).

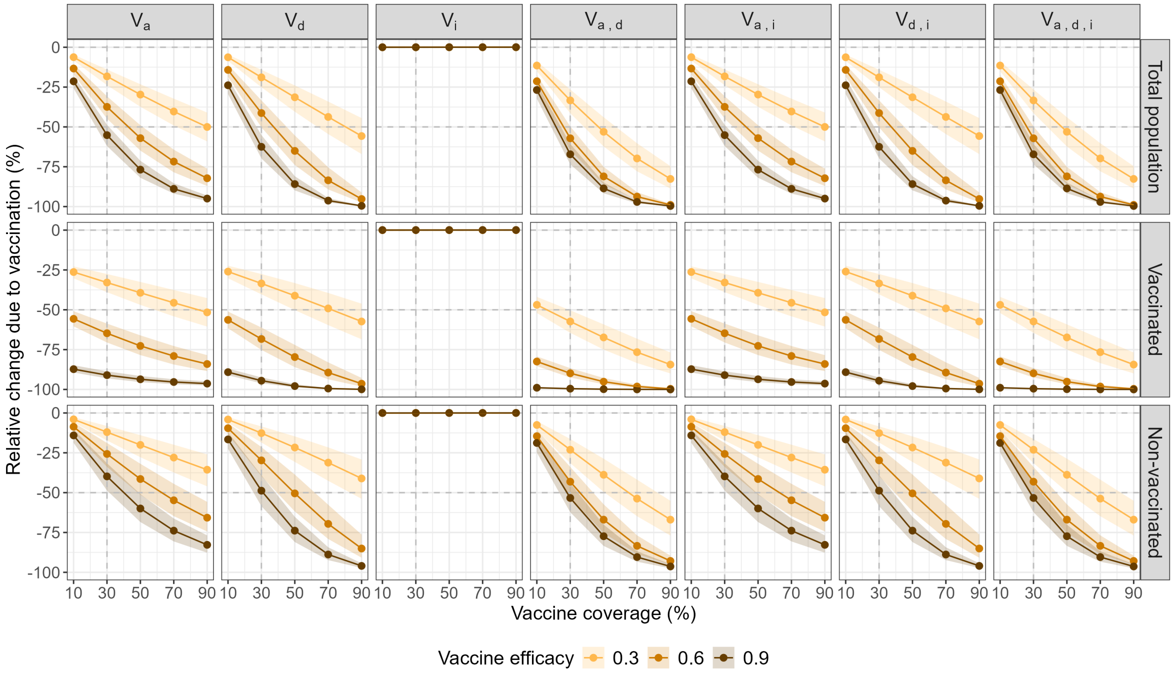

(a) For *S.aureus*

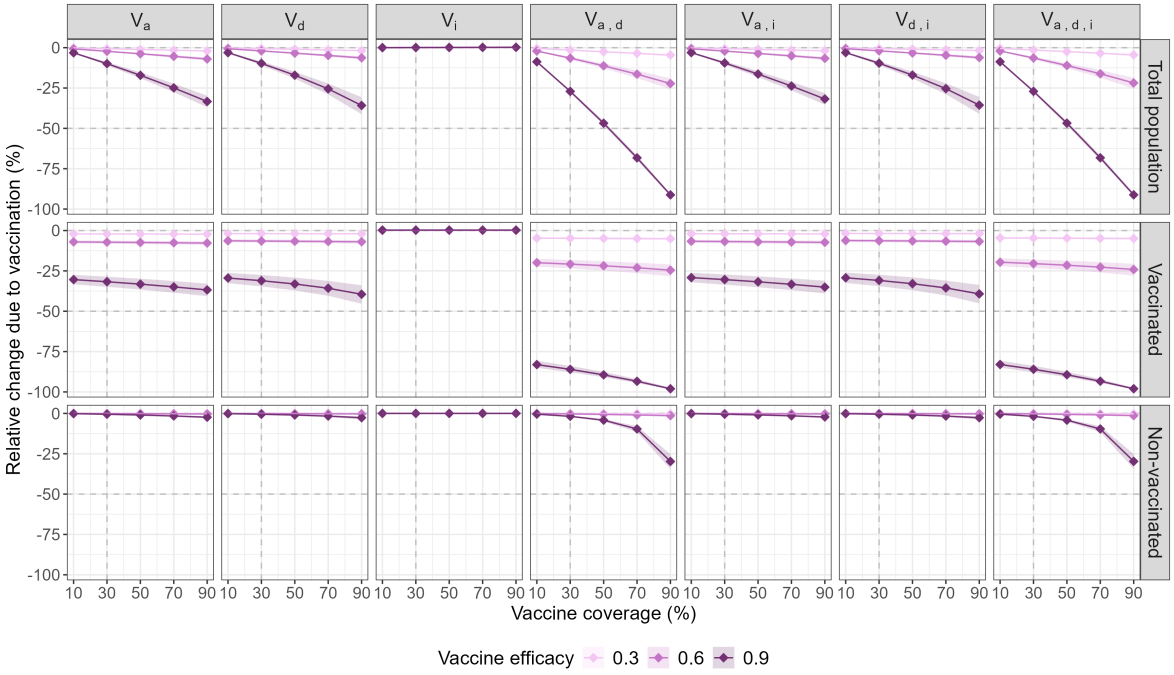

(b) For *E.coli*

Figure A5: **Impact of vaccines on the colonisation prevalence at the end of one year for two bacteria.** The points indicate the median relative reduction in the colonisation prevalence as a function of vaccine coverage, as compared with the no-vaccine prevalence, and the ribbon depicts the 95% interval between the 2.5th and 97.5th percentiles. Each column corresponds to one vaccine type defined in Table 1. For each considered vaccine, three levels of efficacy are considered: 30% (light), 60% (medium) and 90% (dark), assuming identical vaccine-driven impacts on model parameters for vaccines that incorporate several. Each row corresponds to a specific population (total, vaccinated or non-vaccinated).

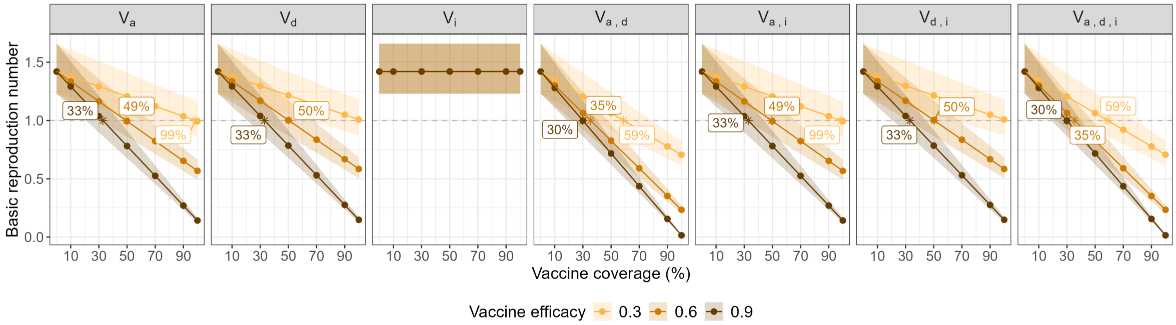

(a) For *S.aureus*

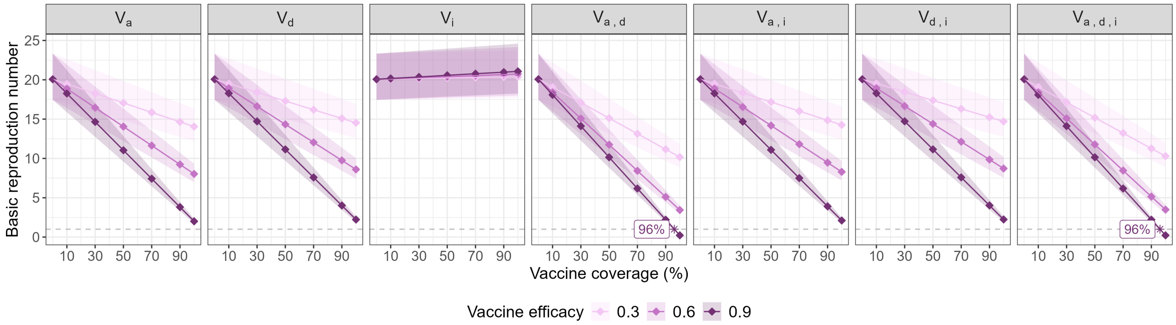

(b) For *E.coli*

Figure A6: **Impact of vaccines on the basic reproduction number for two bacteria.** The points indicate the median basic reproduction number as a function of vaccine coverage, as compared with the no-vaccine reproduction number, and the ribbon depicts the 95% interval between the 2.5th and 97.5th percentiles. Each column corresponds to one vaccine type defined in Table 1. For each considered vaccine, three levels of efficacy are considered: 30% (light), 60% (medium) and 90% (dark), assuming identical vaccine-driven impacts on model parameters for vaccines that incorporate several. The star points are added to represent the limit of $R_0 = 1$. Labels indicate the minimum vaccine coverage needed to have $R_0$ below 1.
